# Alternative epidemic indicators for COVID-19: a model-based assessment of COVID-19 mortality ascertainment in three settings with incomplete death registration systems

**DOI:** 10.1101/2023.01.04.22283691

**Authors:** Ruth McCabe, Charles Whittaker, Richard J. Sheppard, Nada Abdelmagid, Aljaile Ahmed, Israa Zain Alabdeen, Nicholas F. Brazeau, Abd Elhameed Ahmed Abd Elhameed, Abdulla Salem Bin-Ghouth, Arran Hamlet, Rahaf AbuKoura, Gregory Barnsley, James A. Hay, Mervat Alhaffar, Emilie Koum Besson, Semira Mitiku Saje, Binyam Girma Sisay, Seifu Hagos Gebreyesus, Adane Petros Sikamo, Aschalew Worku, Yakob Seman Ahmed, Damen Haile Mariam, Mitike Molla Sisay, Francesco Checchi, Maysoon Dahab, Bilal Shikur Endris, Azra C. Ghani, Patrick G. T. Walker, Christl A. Donnelly, Oliver J. Watson

## Abstract

Not all COVID-19 deaths are officially reported and, particularly in low-income and humanitarian settings the magnitude of such reporting gaps remain sparsely characterised. Alternative data sources, including burial site worker reports, satellite imagery of cemeteries and social-media-conducted surveys of infection, may offer solutions. By merging these data with independently conducted, representative serological studies within a mathematical modelling framework, we aim to better understand the range of under-reporting using the example of three major cities: Addis Ababa (Ethiopia), Aden (Yemen) and Khartoum (Sudan) during 2020. We estimate 69% - 100%, 0.8% - 8.0% and 3.0% - 6.0% of COVID-19 deaths were reported in these three settings, respectively. In future epidemics, and in settings where vital registrations systems are absent or limited, using multiple alternative data sources could provide critically-needed, improved estimates of epidemic impact. However, ultimately, functioning vital registration systems are needed to ensure that, in contrast to COVID-19, the impact of future pandemics or other drivers of mortality are reported and understood worldwide.

**One sentence summary:** We demonstrate the suitability of alternative data sources to assess the under-ascertainment of COVID-19 mortality.

---

Accurate ascertainment of infections, cases and deaths of an emerging infectious disease are vital in order to implement an effective public health response. However, these quantities depend on both testing capacity and robust vital registration systems. Consequently, only a subset of the true burden of a disease is captured within official statistics. During the COVID-19 pandemic, official COVID-19 data obscured the perception of the global burden of the disease (*1*). For example, in some settings with low reported COVID-19 death tolls, serological surveys (serosurveys) have revealed extensive SARS-CoV-2 transmission (*2*–*4*). These levels of transmission are inconsistent with the number of infections expected based on officially reported deaths and estimates of the infection fatality ratio (IFR) (*5, 6*), with the most parsimonious explanation that COVID-19 deaths go unreported (*7*).

Due to the underreporting of COVID-19 deaths (*8*), excess mortality has been used frequently as an alternative means by which to assess the impact of the COVID-19 pandemic (*9*). On 5 May 2022, the World Health Organization (WHO) published estimates of the “full” global death toll of the pandemic based on all-cause excess mortality (*10*). Although not subject to the same biases as COVID-19 deaths, which typically rely on a proven SARS-CoV-2 infection around the time of the death, estimates of excess mortality still require complete data on the total number of deaths from all causes, which in turn requires mechanisms by which to record these deaths. Unfortunately, robust vital registration systems do not exist in many parts of the world, with the WHO estimating in 2020 that 40% of the world’s deaths from all causes occur unregistered (*11*).

In settings without official all-cause mortality data, estimation of excess mortality during the pandemic has relied solely on model-based inference by pooling information from countries with similar socioeconomic and demographic characteristics. An alternative approach is to leverage alternative mortality data sources and epidemic indicators that can rapidly provide a more data-driven understanding of epidemic dynamics, such as social-media-shared obituary notifications (*8*). Characterising the potential biases in these novel data sources is crucial to understanding their suitability for tracking the spread of SARS-CoV-2, as well as more broadly for mortality monitoring in the absence of complete vital registration. In response, we consider here three alternative data sources generated during the pandemic to understand the transmission of SARS-CoV-2 in two cities in sub-Saharan Africa and one in the Middle East. These include burial site worker reports in Addis Ababa (*12*), satellite imagery of cemeteries in Aden (*13*) and social-media-conducted surveys of symptomatic infection in Khartoum (*14*) (Table 1).

**Table 1.** Sources of alternative data collected at study locations and seroprevalence estimates. See Supplementary Material for further details of each study and the specifics of assays used.

| Setting | Alternative data source |  | Seroprevalence |  |  |
| --- | --- | --- | --- | --- | --- |
|  | Overview | Ref | Overview | Reported Estimate | Ref |
| Addis Ababa, Ethiopia | Burial Site Worker Cemetery Reports.<br>01/01/2015 - 26/01/2021 | (12) | Random sample of 956 households.<br>22/07/2020 - 10/08/2020 | Immunoglobulin G (IgG):<br>1.9% (95% CI: 0.4% - 3.7%)<br>Combined IgG/IgM:<br>3.5% (95% CI: 1.7% - 5.4%) | (16) |
| Aden, Yemen | Satellite Imagery of cemetery burials.<br>21/07/2016 - 19/09/2020 | (13) | Cross-sectional household study of 2001 people.<br>28/11/2020 - 13/12/2020 | IgG:<br>25% (95% CI: 23.2% - 26.9%)<br>IgM:<br>0.2% (95% CI: 0.1% - 0.4%)<br>IgG & IgM:<br>2.3% (95% CI: 1.7% - 2.9%)<br>Combined IgG/IgM:<br>27.4% (95% CI: 25.6% - 29.3%) | (17) |
| Khartoum, Sudan | Survey of historic symptomatic infections conducted through social-media channels.<br>26/05/2020 - 03/06/2020 | (14) | Cross-sectional household study of 2375 people.<br>01/03/2021 - 10/04/2021 | Combined IgG/IgM:<br>54.6% (95% CI: 51.4% - 57.8%) | (18) |

In each setting, representative serosurveys have also been conducted independently at different time-points before the availability of vaccines. We incorporate these within a previously developed mathematical modelling framework (*8*) to quantify how informative each data source is for explaining population seroprevalence under different assumptions for the IFR in each setting. In brief, the model uses a Susceptible-Exposed-Infected-Recovered structure, which explicitly captures pathways through hospital care and the impact this has on mortality (see *Supplementary Material* for further information). Our default assumption is that the relationship between the infection fatality ratio (IFR) and age is consistent with the log-linear relationship estimated in high-income settings as estimated in multiple modelling studies (*5, 6*) and meta-analyses (*15*). We test this hypothesis first by fitting to the alternative sources for mortality (Addis Ababa and Aden) and case incidence (Khartoum) data using the relationship between IFR and age as estimated in Brazeau et al. (*5*). From these model fits, we compare the model predicted seroprevalence against the reported seroprevalence estimates using a chi-squared test, arguing that this is good evidence that the alternative datasets are reliable for tracking the transmission of SARS-CoV-2 and the assumed IFR is suitable should these values not differ significantly. Additionally, in settings where the alternative sources and the serosurveys do not agree it presents an opportunity to better understand the biases in these alternative datasets as well as explore alternative hypotheses for the severity of COVID-19 in these settings. Full methods are presented in the Supplementary Material.

## Addis Ababa

In Addis Ababa, excess mortality was derived from cemetery surveillance data collected via the Addis Ababa Mortality Surveillance Program across January 2015 - January 2021, as detailed by Endris et al. (*12*). These data comprise the number of burials per day at all cemeteries in the city according to official records kept by workers at each site. As cremation is not practised in Addis Ababa, this surveillance programme is expected to capture the vast majority of deaths from all causes in the city (*12*).

Reported COVID-19 deaths (*19*) largely matched estimated excess mortality from June 2020 onwards, though this was in part dependent on how baseline mortality was estimated (Figures 1A and 1B). While the annual number of burials in each year from 2015 to 2018 was largely consistent (average 12,862; sd 389), there was a significant reduction in the number of burials observed in 2019 compared to the mean from 2015 - 2018 (total 11,256; chi-squared test p-value <0.001, Figure S3). Therefore, we explored two methods for estimating baseline mortality in 2020, in which we used mortality data either from 2015 to 2019 or exclusively from 2019 (Spearman correlation between excess mortality and COVID-19 deaths over our study period: r = 0.59 (using 2015 to 2019 as baseline) and r = 0.69 (using just 2019 as baseline)). Regardless of the baseline, we observed a peak in excess mortality observed in May 2020 that was not associated with a peak in reported COVID-19 deaths (Figures 1A and 1B). We explored whether this peak could be due to COVID-19 through an additional sensitivity analysis by starting the model from 5 June 2020 instead of 6 April 2020 (the date of the first COVID-19 death) (Figure 1D) (Spearman correlation between excess mortality and COVID-19 deaths from June 2020 onwards: r = 0.74 and r = 0.75 for 2015-to-2019 and 2019-only baselines, respectively). Therefore, we considered four excess mortality scenarios in total for Addis Ababa, which we assumed to be equally plausible a priori.

**Figure 1:**
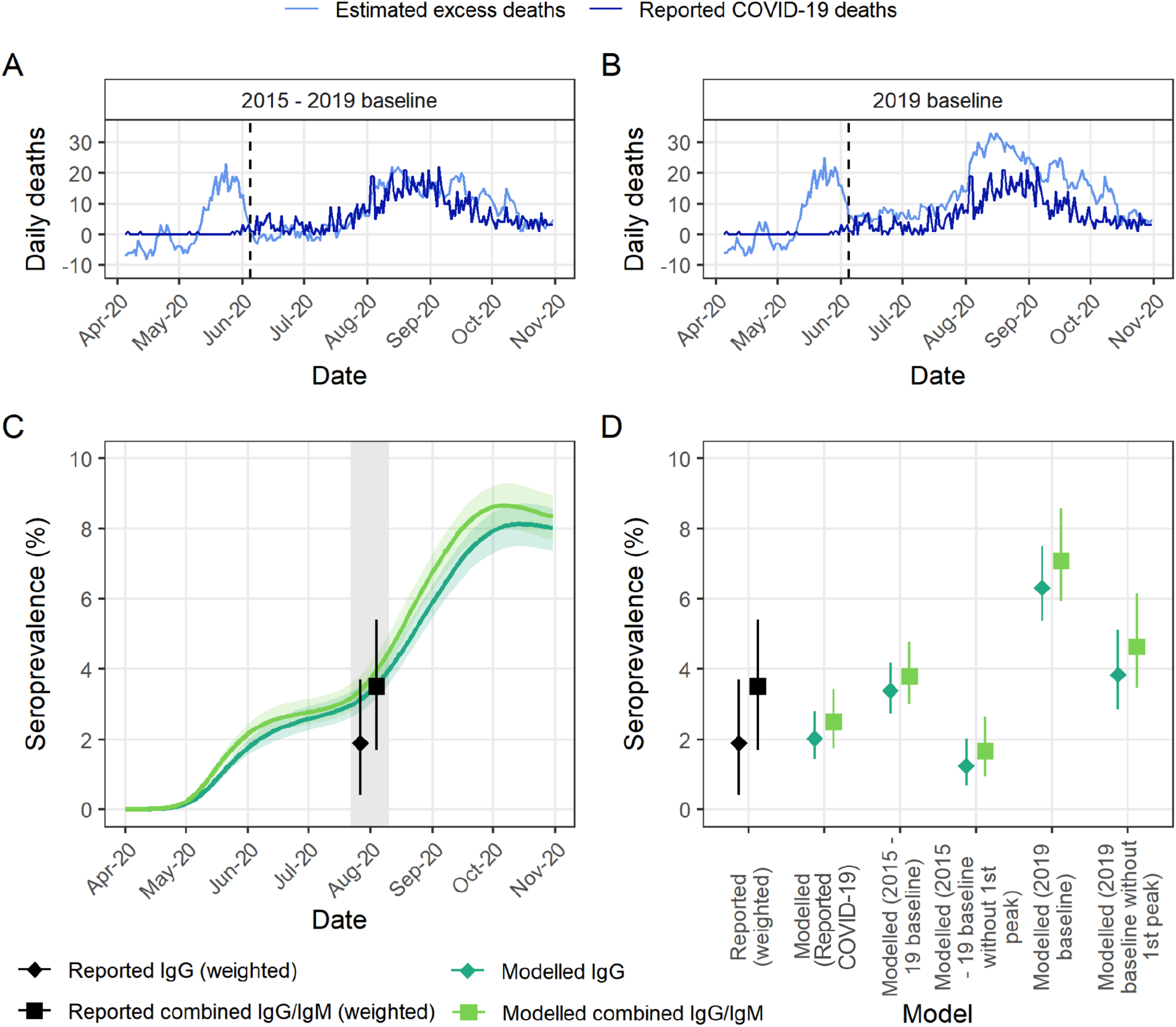
Mortality reporting and seroprevalence in Addis Ababa, Ethiopia in 2020. **(A)** Reported COVID-19 deaths compiled from the Ethiopian Public Health Institute and estimated excess mortality using cemetery surveillance from (*12*) using data from 2015 - 2019 to derive the baseline. Dashed line indicates the end of the early peak in excess mortality featured in the sensitivity analysis. **(B)** As in **(A)**, but with excess mortality derived from 2019 data only. **(C)** Estimated seroprevalence from model fit to excess mortality under 2015 - 2019 baseline (median and 95% credible intervals) with the first peak compared to reported values by Abdella et al. (*16*). Grey shaded area highlights the sampling period of the serosurvey, with weighted reported estimates both corresponding to this entire period. **(D)** Seroprevalence estimated under models fit to either reported COVID-19 or different estimates of excess mortality with different baselines (median and 95% credible intervals) compared to reported seroprevalence in (*16*).

We fitted the mathematical model separately to reported COVID-19 deaths and the four estimated excess mortality time series and compared the number of infections estimated by the model to those at the time of a serosurvey conducted by Abdella et al. (*16*) in July - August 2020. The study used random sampling of 956 households in the city and reported a weighted prevalence of Immunoglobulin G (IgG) and combined IgG/IgM antibodies of 1.9% (0.4% - 3.7%) and 3.5% (1.7% - 5.4%), respectively, having post-stratified by age and sex to account for potential biases in their sample (see Supplementary Material for further information).

The model fit to COVID-19 deaths produced estimates of seroprevalence not statistically different from the values reported by Abdella et al. (*16*) (p-values: 0.884 and 0.334 for IgG and combined IgG/IgM, respectively), suggesting that officially reported deaths are representative of the true COVID-19 death toll in this setting. Similarly, we observed that the majority of the seroprevalence estimates based on model fits to the excess mortality inferred from cemetery burial data were not statistically significantly different from the reported seroprevalence in Abdella et al. for both antibody types (Table S2). However, this is unsurprising given the similarity in the COVID-19 and excess mortality time series (Figures 1A and 1B). The exception to this was the model fit to excess mortality inferred using 2019 only as the baseline and including the first peak (p-values: <0.001 and 0.002 for IgG and combined IgG/IgM antibodies, respectively), with the resulting model-estimated seroprevalence three times and twice as great as the reported prevalence of IgG and combined IgG/IgM antibodies of Abdella et al. (Figure 1D). This suggests that excess mortality under this baseline scenario overestimated COVID-19 mortality and is unsuitable for informing the size of the COVID-19 epidemic. Consequently, we approximate that, depending on the selection of mortality baseline, between 68.7% (1064/1549, reflecting the 2015 - 2019 scenario with the first peak in May included) - 100% of COVID-19 deaths were reported across April - November 2020 in Addis Ababa. The upper estimate of 100% (complete) reporting reflects both the near agreement between the cemetery-inferred excess mortality and reported COVID-19 deaths, and the non-significant difference between the seroprevalence inferred based on model fits to reported COVID-19 deaths and the reported seroprevalence (Figure 1C, Table S5).

### Aden

In Aden, excess mortality was estimated from satellite imagery surveillance of all cemeteries in the city to estimate daily burials between July 2016 - September 2020, by quantifying expansions to cemetery surface area and validating this with civil death registrations (*13*). From these data, a wave of excess mortality was estimated to have occurred between April - September 2020, peaking in early June. Reported COVID-19 deaths were only available via scraping death counts tweeted daily by the Yemen Supreme National Emergency Committee for COVID-19 (*20*). Excess mortality estimates suggested 2120 (95% CI: 424 - 4137) excess deaths occurred across 1st April - 19 September 2020, compared to just 34 officially reported COVID-19 deaths during this same period (*20*) implying substantial under-ascertainment of COVID-19 mortality (Figure 2A).

**Figure 2:**
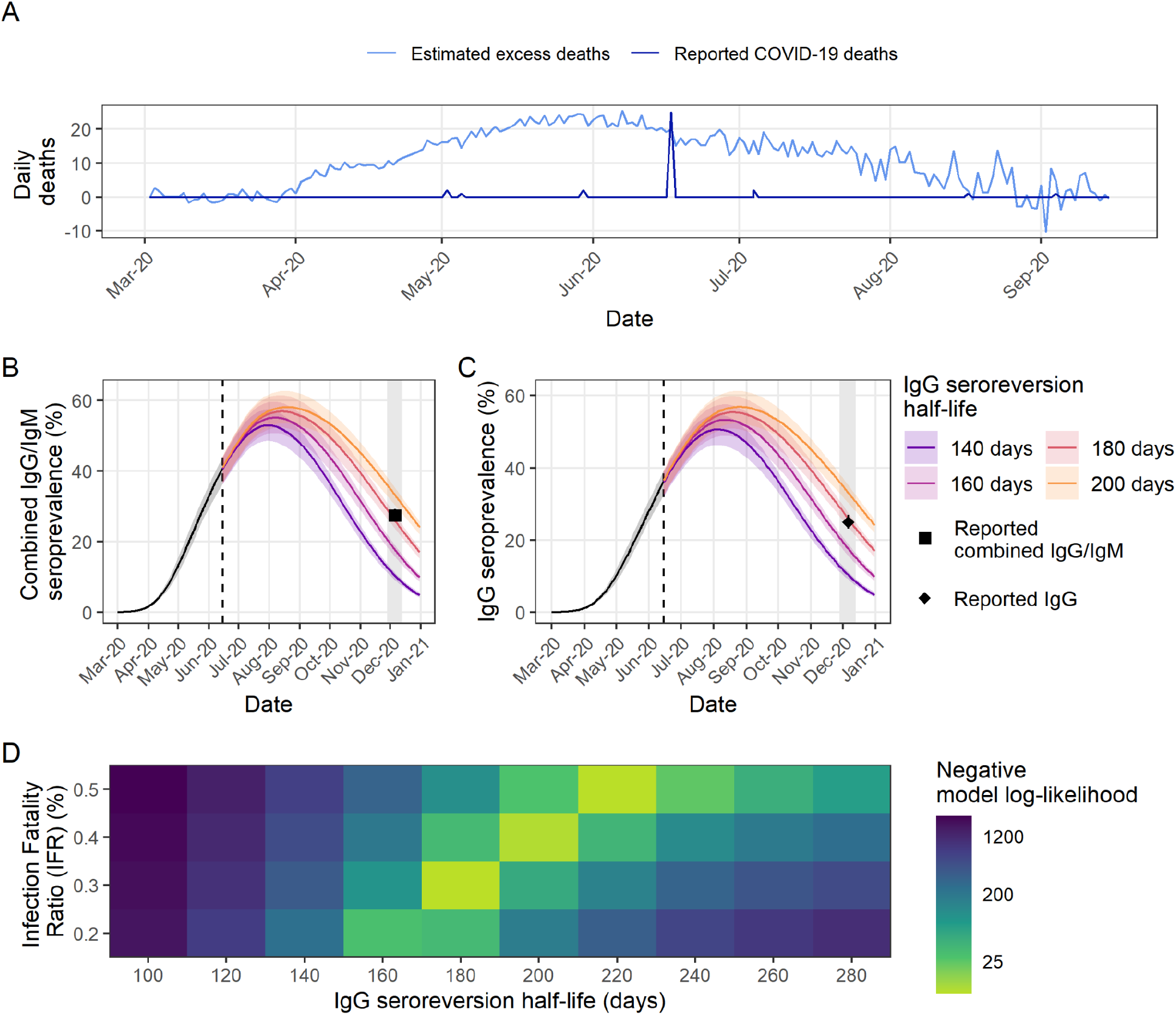
Mortality reporting and seroprevalence in Aden, Yemen in 2020. **(A)** Reported COVID-19 deaths and estimated excess mortality from satellite surveillance of burials from (*13*). **(B)** Estimated seroprevalence of combined IgG/IgM (positive for either IgG and/or IgM) from the model fitted to excess mortality using the default Infection Fatality Ratio (IFR = 0.3%) under different IgG seroreversion half-lives compared to observed values from (*17*) (reported seroprevalence: 27.4% (95% CI 25.6% - 29.3%)). Dashed vertical line indicates point in time when seroprevalence dynamics divert due to difference in IgG half life modelled. **(C)** As in **(B)** but for IgG antibodies (reported seroprevalence: 25.0% (95% CI 23.2% - 26.9%)). **(D)** Combined negative log-likelihood of models of IgG and combined IgG/IgM antibodies under varying assumptions of the IFR and IgG seroreversion half-life. In **(B)** and **(D)** the IgM seroreversion half-life is held constant at 50 days. Values associated with the highest log-likelihoods (shown here in light-green) indicate the best fit of the parameter values to the observed data.

In December 2020, Bin-Ghouth et al. (*17*) conducted a cross-sectional household serosurvey of 2001 people and estimated that 25.0% (95% CI: 23.2% - 26.9%) and 0.2% (95% CI: 0.1% - 0.4%) of the population had either IgG or IgM antibodies to SARS-CoV-2, respectively, with a total of 27.4% (95% CI: 25.6% - 29.3%) estimated to have IgG and/or IgM antibodies (combined IgG/IgM). Due to the long delay of approximately five months between the peak in excess mortality and the implementation of the serosurvey in this setting, it was necessary to consider the decay of antibodies after infection over time. Antibody kinetics vary substantially among individuals resulting in high uncertainty around the duration of seropositivity after infection (*21*). To the best of our knowledge, these parameters had not been quantified for the assay used in this study and so we conducted sensitivity analyses to explore the impact of different assumptions for IgG and IgM seroreversion. IgM antibodies endure for a substantially shorter duration than IgG antibodies. As such, we found that varying the IgM half-life had a minimal impact in the context of this analysis and thus assumed a central value of 50 days as in (*5*) (statistically insignificant differences with reported values, Figure S11, Table S3), but that the IgG half-life was an important determinant of our estimated seroprevalence (Figure 2, Table S3).

Seroprevalence inferred from the model fit to reported COVID-19 deaths was statistically significantly different from that reported for all antibody types and under all seroreversion half-lives considered (p-values: <0.001 for IgG and combined IgG/IgM, <0.009 for IgM), suggesting that official reported COVID-19 deaths substantially underestimate the true death toll. However, in model fits to the satellite-derived excess mortality, we found no statistically significant differences between reported and modelled seroprevalence of IgG and combined IgG/IgM antibodies (p-values: 0.651 and 0.563, respectively) with an assumed IgG seroreversion half-life of 180 days. This finding suggests that these data are a more accurate representation of the first COVID-19 outbreak in Aden than officially reported deaths.

Although the default IFR (IFR = 0.3%) and IgG seroreversion half-life (180 days) maximised the combined model log-likelihood of IgG and combined IgG/IgM antibodies, we were able to find other parameter combinations which could also explain the reported seroprevalence (Figure 2). We found that a slightly higher IFR of 0.4% or 0.5% combined with a slightly longer seroreversion half-life of 200 or 220 days, respectively, also produced non-statistically significant differences between reported and modelled seroprevalence of both antibody types (p-values: 0.289 and 0.649 for IgG and 0.944 and 0.385 for combined IgG/IgM antibodies, respectively) and produced similarly high log-likelihoods (Figure 2, Table S3). Although a lower IFR of 0.2% was able to also produce non-statistically significant differences when using shorter seroreversion half-lives (Table S3), the model was unable to recreate the excess deaths from the satellite data, suggesting that IFR in Aden was greater than 0.2% (Figure S10).

Overall, the broad agreement between the two independent data sources under plausible assumptions of antibody seroreversion and the IFR strongly supports COVID-19 to be the driver of the observed satellite-inferred excess mortality. Consequently, we estimate that just 1.6% (34/2120) of COVID-19 deaths in Aden in summer 2020 were captured in official statistics (Table S5). Based on the uncertainty in the satellite-inferred excess mortality (95% CI: 424 - 4137), we report our uncertainty in the reporting fraction to be between 0.8% - 8.0%.

### Khartoum

In Khartoum, we leveraged an online survey distributed through social media channels (*14*). The survey presented respondents with a list of symptoms known to be associated with SARS-CoV-2 infection and used to triage patients in Sudan. Respondents were asked which of these symptoms they had experienced since the start of the pandemic. Respondents were also asked both if they had received i) a SARS-CoV-2 diagnostic test and ii) the outcome of any test taken. From this survey, the SARS-CoV-2 infection status of survey participants who had not received a COVID-19 test was inferred based on their reported symptom(s), resulting in an estimate of the symptomatic attack rate in the general population. The survey collected responses opportunistically, resulting in 5,018 responses from individuals over 15 years old throughout Khartoum State between 26 May and 3 June 2020. From these respondents, 11.0% of individuals were estimated to have experienced a symptomatic infection by 3 June, with lower and upper estimates (based on different methods for inferring symptomatic infection) of 8.3% and 13.7%.

By the end of March 2021 (prior to a pause in publication of COVID-19 reports by the Sudan Federal Ministry of Health), 938 COVID-19 deaths had been reported in Khartoum (Figure 3A). We fit the transmission model to the observed COVID-19 deaths making the assumption that the reported COVID-19 deaths reflected a fixed proportion of the true total number of COVID-19 deaths over time (Figure S12, S13). From these, our central estimate is that 4% of COVID-19 deaths were reported based on comparison against the 11.0% of individuals estimated to have experienced a symptomatic infection by 3 June (Figure 3B). We report our uncertainty in the reporting fraction to be between 3% - 6%, with the modelled 95% confidence interval for the cumulative proportion of symptomatic infections overlapping with the lower or upper bounds reported of 8.3% and 13.7% (Figure 3B).

**Figure 3.**
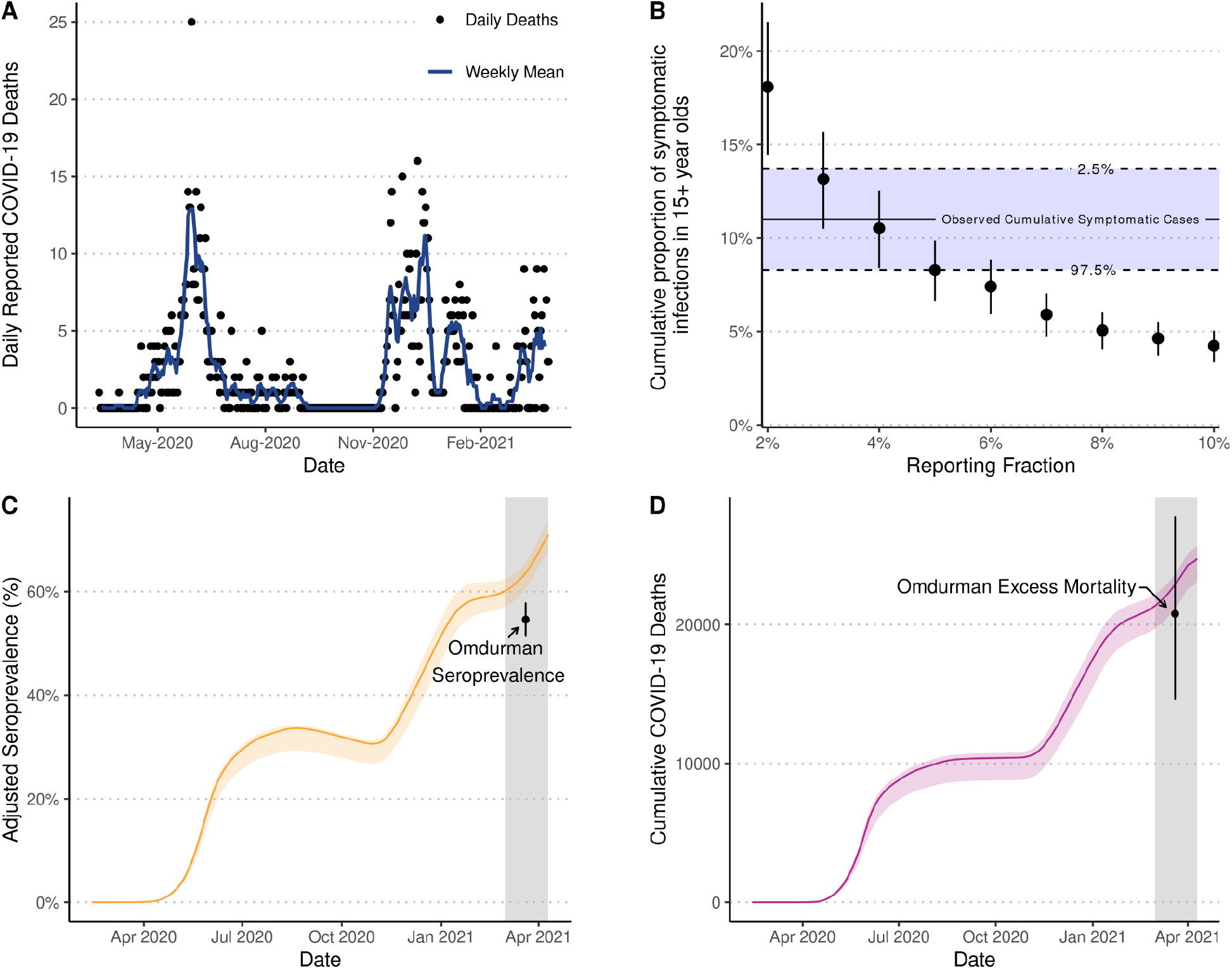
Estimates of under-ascertainment of deaths in Khartoum. **(A)** Daily and weekly mean reported COVID-19 deaths in Khartoum. We fit models to the reported COVID-19 deaths in **(A)** under different assumptions for what proportion of the true number of COVID-19 deaths these represent (reporting fractions). In **(B)** the resultant model fits were used to estimate the proportion of individuals aged over 15 years that would have experienced a symptomatic COVID-19 infection by 3 June 2020. The points and vertical bars show the median and 95% CI for each model fit and are compared against the observed cumulative number of symptomatic cases in Khartoum estimated from a social-media-distributed survey (*14*), suggesting that 4% of COVID-19 deaths were detected. A cross-sectional, household mortality and serosurvey conducted in Omdurman in March - April 2021 estimated seroprevalence to be 54.6% (95% CI: 51.4% - 57.8%) after adjusting for specific test performance (*18*). This estimate is depicted in **(C)** by point and whiskers and is compared against the adjusted seroprevalence (median and 95% CI shown in orange) predicted by a model fit with an assumed mortality reporting fraction of 4%. Based on the same survey, 20,766 (95% CI: 14,641 - 27,750) excess deaths are estimated to have occurred across Khartoum state. This estimate is depicted in **(D)** by point and whiskers and is compared against the cumulative number of COVID-19 deaths (median and 95% CI shown in purple) predicted by a model fit with an assumed mortality reporting fraction of 4%. In **(C)** and **(D)** the grey shaded area highlights the sampling period of the serosurvey and mortality survey in Omdurman.

Between March 1 and April 10 2021, a cross-sectional household mortality and seroprevalence survey was conducted in Omdurman - a city in Khartoum state estimated to encompass ~35% of the population of Khartoum state (*18*). In this survey, 54.6% (95% CI: 51.4% - 57.8%) of the population were seropositive and 7,113 excess deaths (95% CI: 5,015 - 9,505) were estimated to have occurred, which when scaled linearly to reflect the population of Khartoum state would be 20,766 (95% CI: 14,641 - 27,750) excess deaths (*18*). By comparison, with 4% of COVID-19 deaths assumed to have been detected, we estimate that 63.6% (95% CI: 60.6% - 65.7%) of the population would have been seropositive by March 20 2021 (Figure 3C), which is significantly different to the reported seroprevalence of 54.6% (chi-squared test p-value <0.001, Table S4), and that 22,870 (95% CI: 21,150 - 23,640) deaths would have occurred (Figure 3D). Consequently, while the inferred reporting fraction of 4% based on the symptomatic survey yields estimates of the epidemic size that are similar to those observed in Omdurman almost a year later, estimates of the epidemic size are most accurately recreated with an assumed reporting fraction of 4.5% (Figure S14, Table S5).

## Conclusions

In this study, we demonstrate the validity of using alternative data sources, namely burial site worker reports in Addis Ababa (*12*), satellite imagery of cemeteries in Aden (*13*) and social-media-conducted surveys of symptomatic infection in Khartoum (*14*), to track the dynamics and burden of the COVID-19 epidemic in each of these settings. By incorporating these data within a previously-published mathematical modelling framework (*8*), we could infer the number of infections implied by each source and validate the accuracy of these results by comparing them with independently-conducted serosurveys. Consequently, we estimate that between 67% - 100% of COVID-19 deaths were reported in Addis Ababa, while in Aden and Khartoum, we demonstrated that there are likely to have been large unobserved epidemics with 0.8% - 8.0% and 3.0% - 6.0% of deaths reported, respectively (Table S5).

Understanding the impact and burden of an infectious disease is integral to facilitating an effective public health response. Our results challenge the perception of the global burden of the disease as reported in official figures which, as shown here in the cases of Aden and Khartoum, can suffer from under-reporting in resource-poor settings. In each setting, we found statistically insignificant differences between our model-estimated seroprevalence based on the alternative data source and those reported in serosurvey, each of which tested at least 950 participants. This provides reasonable confidence in the accuracy of these sources to describe and understand the respective epidemic dynamics (Tables S2 - S4). As such, we consider that these alternative data sources represent a viable, and likely cost-effective, route to estimating disease burden and impact in settings where high-quality civil registration system mortality data are not currently available.

Our framework utilises the IFR when comparing mortality inferred from alternative data sources to reported seroprevalence. Although this parameter has not been estimated directly within the three settings in this study, we have demonstrated that the default IFR accounting for the demography in Addis Ababa (IFR = 0.22%) estimated by Brazeau et al. (*5*) accurately captured the reported seroprevalence. However, in Aden, our results suggested that the IFR is at least 0.3%, which is higher than the IFR estimated by Brazeau et al. after accounting for the demography in Aden (IFR = 0.17%). Similarly, in Khartoum, an IFR of approximately 0.38% accurately captured the reported seroprevalence, which is notably higher than the default IFR estimated by Brazeau et al. after accounting for the demography in Khartoum (IFR = 0.20%). Importantly, these studies should not be viewed as an accurate assessment of the IFR, with the mortality inferred from these alternative data unlikely to perfectly reflect the true COVID-19 mortality in each setting. Nonetheless, in addition to demonstrating these data are indeed informative for describing and understanding epidemic dynamics, our results provide further evidence that the relationship between age and IFR that has been consistently observed in high-income countries (*6, 22*) also exists in these low-income settings. It is further possible that the aforementioned relationship with age could lead to an underestimation of the IFR in some settings, as suggested by a recent meta-analysis of data from developing countries (*7*) and supported by our presented analyses of data from Aden and Khartoum.

Our results could be further corroborated via sources other than those directly used in our analyses. In Aden, multiple reports of hospital capacity being reached coincided with the epidemic peak estimated via satellite image-derived excess mortality (*13, 23*). The saturation of healthcare facilities could also explain the higher IFR indicated in this setting. A similar situation may also have occurred in Khartoum, with over 70% of health centres closed in Khartoum as a COVID-19 containment measure during the first wave (*24*). Additional serosurveys have also been conducted in each setting. In Khartoum, a seroprevalence survey conducted in 22 neighbourhoods and relying on voluntary enrollment organised through Resistance Committees estimated a seroprevalence of 18.3% (95% CI: 16.0% - 20.9%) using rapid antibody immunochromatography tests between May 22 - July 5 2020 (*25*). PCR-confirmed prevalence of infection in the same survey was estimated at 35.0% (95% CI: 32.1% - 38.0%), raising concerns that the sampling scheme had resulted in an upward bias. However, the seroprevalence inferred from our model fits with an assumed reporting fraction of 4% was consistent with this survey (p-value = 0.493), confirming previous viral kinetics modelling of this study (*26*). In Addis Ababa, higher seroprevalence of combined IgG/IgM antibodies has been estimated in other surveys (*27*), which estimated a value of 10.9% in August 2020, with this value rising to 53.7% in December 2020. However, these estimates were derived from sampling focused on healthcare workers, who are known to have a higher risk of exposure to SARS-CoV-2 than the general population (*28*). Similarly, samples collected from Médecins Sans Frontières staff in Aden between September - November 2020 and tested using rapid serology lateral flow tests resulted in an estimated seroprevalence of 19.4% (95% CI: 17.9% – 20.8%) (*29*). However, a follow-up survey of Médecins Sans Frontières staff in Aden during January 2021 (after a period with very few COVID-19 cases reported in Aden (*20*)) but analysed using an electrochemiluminescence immunoassay resulted in an estimated seroprevalence of 59.0% (95% CI: 52.2% – 65.9%) (*29*). This study both shows high exposure in healthcare staff but also demonstrates the importance of accounting for waning rapid test sensitivity. Lastly, excess mortality published by the WHO (*10*) yielded similar magnitudes to those estimated by our analyses in Aden and Khartoum. However, we estimate a much greater reporting fraction in Addis Ababa than the corresponding national estimate (Table S5, S6 and Figure S15), highlighting how death registration measured in individual cities or locations often will not reflect the realities of the whole countries, especially during periods of crises.

There are a number of limitations of this analysis. First, our estimated reporting fractions correspond to the entire study period and do not account for time-varying trends in detection of COVID-19 deaths. Second, there is uncertainty as to whether all excess deaths can be attributed directly due to COVID-19, which is why we have conducted extensive sensitivity analyses to assess the robustness of our analysis in all three settings. Third, SeroTracker (*30*) suggested all three serosurveys may be subject to ‘moderate’ bias, with corresponding estimates defined as “likely correct for the target population”, as opposed to “very likely correct” for studies with ‘low’ bias, based on nine criteria including sampling technique, sample size and consistency of reporting results (*30, 31*). Nonetheless, throughout our study we have considered the full range of uncertainty estimated in the serosurveys and also estimated from our model fits, with these intervals shown to overlap under certain assumptions in each study. Analogously to the mortality data, we have focussed on comparing the range of uncertainty intervals as opposed to point estimates alone. This is also applicable to the social media-based survey in Khartoum, which may suffer from self-reporting biases. Fourth, SARS-CoV-2 assays are often developed on high-income populations and the applicability to other settings has been questioned (*32*), which could influence the results. Fifth, we were unable to obtain specific seroconversion and seroreversion rate estimates for the assays used in these studies and had to rely on a sensitivity analysis around estimates derived from studies of other assays. Similarly, our modelling framework operates under the assumption of independence between the likelihood of the detection of IgG and combined IgG/IgM antibodies, due to a lack of data to prove otherwise. These assumptions may explain why we were unable to recreate the relatively large difference between IgG and combined IgG/IgM antibodies observed in the serosurvey in Addis Ababa, despite adjusting for different seroreversion half-lives of these two antibody measurements. Crucially, our qualitative conclusions about the significant degree of underreporting and the usefulness of these alternative data sources are robust to the limitations described above. Consequently, while there are likely biases in the alternative data sources leveraged, we argue that these biases are substantially smaller than those in official reported COVID-19 statistics and are thus vitally useful as proxies for COVID-19 mortality and epidemic dynamics.

The COVID-19 pandemic has caused a substantial loss of life, but reported deaths are likely to only capture a small proportion of the true death toll in settings without robust vital registration. In the absence of all-cause mortality data, we have validated the suitability of alternative data sources, namely burial site worker reports, satellite imagery of cemeteries and social-media-conducted surveys of symptomatic infection, in tracking epidemics in Addis Ababa, Aden and Khartoum across 2020. These sources provide a critical insight into the dynamics of SARS-CoV-2 in these settings and contradict the hypothesis that low-income countries were spared the worst of the pandemic. The global community must prioritise providing support and investment in the development of vital registration systems to capture all-cause mortality across the globe. Better vital registration is not only essential for future pandemic preparedness plans but also is foundational for evaluating progress made across multiple areas of public health and is essential for the Sustainable Development Goals’ mission to “leave no-one behind” (*33*).

## Data Availability

All data and code are provided at https://github.com/mrc-ide/covid-alternative-mortality/

## Funding

This work was supported by the NIHR HPRU in Emerging and Zoonotic Infections, a partnership between PHE, University of Oxford, University of Liverpool and Liverpool School of Tropical Medicine [grant number NIHR200907 supporting RM and CAD]; and the MRC Centre for Global Infectious Disease Analysis [grant number MR/R015600/1], which is jointly funded by the UK Medical Research Council (MRC) and the UK Foreign, Commonwealth & Development Office (FCDO), under the MRC/FCDO Concordat agreement and is also part of the EDCTP2 programme supported by the European Union (EU). OJW is supported by a Schmidt Science Fellowship in partnership with the Rhodes Trust. This publication is supported by the Centers for Disease Control and Prevention of the U.S. Department of Health and Human Services (HHS) as part of financial assistance award U01GH002319. The contents are those of the author(s) and do not necessarily represent the official views of, nor an endorsement, by CDC/HHS, or the U.S. Government

### Disclaimer

“The views expressed are those of the authors and not necessarily those of the United Kingdom (UK) Department of Health and Social Care, EU, FCDO, MRC, National Health Service, NIHR, or PHE. The funding bodies had no role in the design of the study, analysis and interpretation of data and in writing the manuscript.”

## Author contributions

OJW conceived the idea for the study and designed the methodology used. RM, CW and OJW conducted the analysis with contributions to software and model fitting by RS, NB, GB, AH and PGTW. NA, AA, AEAAE, RAK, IZA and MD contributed data, model parameters and situational awareness for the analysis in Khartoum. BA MA, ASBG, EKB and FC contributed data, model parameters and situational awareness for the analysis for the Aden study. SMJ, BGS, SHG, AP, AW, YS, DHM, MMS, AK and BSE contributed data, model parameters and situational awareness for the analysis for the Addis Ababa study. ACG and CAD provided supervision throughout. RM and OJW wrote the first draft of the manuscript. All authors contributed to redrafting.

## Data and materials availability

All data and code are provided at https://github.com/mrc-ide/covid-alternative-mortality/ (*34*).

## List of Supplementary Materials

Materials and Methods

Tables S1 to S6

Figures S1 to S15

## References

1. D. Ghosh, J. A. Bernstein, T. B. Mersha, COVID-19 pandemic: The African paradox. J. Glob. Health. 10, 020348 (2020).

2. S. Uyoga, I. M. O. Adetifa, H. K. Karanja, J. Nyagwange, J. Tuju, P. Wanjiku, R. Aman, M. Mwangangi, P. Amoth, K. Kasera, W. Ng’ang’a, C. Rombo, C. Yegon, K. Kithi, E. Odhiambo, T. Rotich, I. Orgut, S. Kihara, M. Otiende, C. Bottomley, Z. N. Mupe, E. W. Kagucia, K. E. Gallagher, A. Etyang, S. Voller, J. N. Gitonga, D. Mugo, C. N. Agoti, E. Otieno, L. Ndwiga, T. Lambe, D. Wright, E. Barasa, B. Tsofa, P. Bejon, L. I. Ochola-Oyier, A. Agweyu, J. A. G. Scott, G. M. Warimwe, Seroprevalence of anti–SARS-CoV-2 IgG antibodies in Kenyan blood donors. Science. 371, 79–82 (2021).

3. K. Wiens, P. N. Mawien, J. Rumunu, D. Slater, F. Jones, S. Moheed, A. Caflisch, B. Bior, I. Jacob, R. L. Lako, A. G. Guyo, O. O. Olu, S. Maleghemi, A. Baguma, J. J. Hassen, S. Baya, L. Deng, J. Lessler, M. Demby, V. Sanchez, R. Mills, C. Fraser, R. Charles, J. Harris, A. Azman, J. Wamala, Seroprevalence of Severe Acute Respiratory Syndrome Coronavirus 2 IgG in Juba, South Sudan, 2020. Emerging Infectious Disease journal. 27, 1598 (2021).

4. H. C. Lewis, H. Ware, M. Whelan, L. Subissi, Z. Li, X. Ma, A. Nardone, M. Valenciano, B. Cheng, K. Noel, C. Cao, M. Yanes-Lane, B. Herring, A. Talisuna, N. Nsenga, T. Balde, D. A. Clifton, M. Van Kerkhove, D. L. Buckeridge, N. Bobrovitz, J. Okeibunor, R. K. Arora, I. Bergeri, the UNITY Studies Collaborator Group, SARS-CoV-2 infection in Africa: A systematic review and meta-analysis of standardised seroprevalence studies, from January 2020 to December 2021. bioRxiv (2022),, doi:10.1101/2022.02.14.22270934.

5. N. Brazeau, R. Verity, S. Jenks, H. Fu, C. Whittaker, P. Winskill, I. Dorigatti, P. Walker, S. Riley, R. P. Schnekenberg, Others, Report 34: COVID-19 infection fatality ratio: estimates from seroprevalence, doi:10.25561/83545.

6. M. O’Driscoll, G. Ribeiro Dos Santos, L. Wang, D. A. T. Cummings, A. S. Azman, J. Paireau, A. Fontanet, S. Cauchemez, H. Salje, Age-specific mortality and immunity patterns of SARS-CoV-2. Nature. 590, 140–145 (2021).

7. A. T. Levin, N. Owusu-Boaitey, S. Pugh, B. K. Fosdick, A. B. Zwi, A. Malani, S. Soman, L. Besançon, I. Kashnitsky, S. Ganesh, A. McLaughlin, G. Song, R. Uhm, D. Herrera-Esposito, G. de los Campos, A. C. P. Antonio, E. B. Tadese, G. Meyerowitz-Katz, Assessing the burden of COVID-19 in developing countries: systematic review, meta-analysis and public policy implications. BMJ Global Health. 7, e008477 (2022).

8. O. J. Watson, M. Alhaffar, Z. Mehchy, C. Whittaker, Z. Akil, N. F. Brazeau, G. Cuomo-Dannenburg, A. Hamlet, H. A. Thompson, M. Baguelin, R. G. FitzJohn, E. Knock, J. Lees, L. K. Whittles, T. Mellan, P. Winskill, Imperial College COVID-19 Response Team, N. Howard, H. Clapham, F. Checchi, N. Ferguson, A. Ghani, E. Beals, P. Walker, Leveraging community mortality indicators to infer COVID-19 mortality and transmission dynamics in Damascus, Syria. Nat. Commun. 12, 2394 (2021).

9. D. M. Weinberger, J. Chen, T. Cohen, F. W. Crawford, F. Mostashari, D. Olson, V. E. Pitzer, N. G. Reich, M. Russi, L. Simonsen, A. Watkins, C. Viboud, Estimation of Excess Deaths Associated With the COVID-19 Pandemic in the United States, March to May 2020. JAMA Intern. Med. 180, 1336–1344 (2020).

10. Estimates of Excess Mortality Associated With COVID-19 Pandemic (as of 25 March 2022). Geneva: World Health Organization, 2022 (2022), (available at https://www.who.int/data/stories/global-excess-deaths-associated-with-covid-19-january-20 20-december-2021).

11. World Health Organization, SCORE global report 2020, (available at https://www.who.int/data/stories/score-global-report-2020---a-visual-summary).

12. B. S. Endris, S. M. Saje, Z. T. Metaferia, B. G. Sisay, T. Afework, Y. G. Mengistu, E. H. Fenta, S. H. Gebreyesus, A. Petros, A. Worku, Y. Seman, D. H. Mariam, M. M. Sisay, A. Karim, Excess Mortality in the Face of COVID-19: Evidence from Addis Ababa Mortality Surveillance Program (2021),, doi:10.2139/ssrn.3787447.

13. E. S. Koum Besson, A. Norris, A. S. Bin Ghouth, T. Freemantle, M. Alhaffar, Y. Vazquez, C. Reeve, P. J. Curran, F. Checchi, Excess mortality during the COVID-19 pandemic: a geospatial and statistical analysis in Aden governorate, Yemen. BMJ Glob Health. 6 (2021), doi:10.1136/bmjgh-2020-004564.

14. F. Mohamed, S. S. Satti, Maram, A. Ahmed, S. F. Babiker, Estimation of Coronavirus (COVID-19) Infections in Khartoum State (Sudan). ResearchGate (2020) (available at https://www.researchgate.net/publication/342122988_Estimation_of_Coronavirus_COVID-19_Infections_in_Khartoum_State_Sudan).

15. A. T. Levin, W. P. Hanage, N. Owusu-Boaitey, K. B. Cochran, S. P. Walsh, G. Meyerowitz-Katz, Assessing the age specificity of infection fatality rates for COVID-19: systematic review, meta-analysis, and public policy implications. Eur. J. Epidemiol. 35, 1123–1138 (2020).

16. S. Abdella, S. Riou, M. Tessema, A. Assefa, A. Seifu, A. Blachman, A. Abera, N. Moreno, F. Irarrazaval, G. Tollera, D. Browning, G. Tasew, Prevalence of SARS-CoV-2 in urban and rural Ethiopia: Randomized household serosurveys reveal level of spread during the first wave of the pandemic. EClinicalMedicine. 35, 100880 (2021).

17. A. S. Bin-Ghouth, S. Al-Shoteri, N. Mahmoud, A. Musani, N. M. Baoom, A. A. Al-Waleedi, E. Buliva, E. A. Aly, J. D. Naiene, R. Crestani, M. Senga, A. Barakat, L. Al-Ariqi, K. Z. Al-Sakkaf, A. Shaef, N. Thabit, A. Murshed, S. Omara, SARS-CoV-2 seroprevalence in Aden, Yemen: a population-based study. Int. J. Infect. Dis. 115, 239–244 (2022).

18. W. Moser, M. A. H. Fahal, E. Abualas, S. Bedri, M. T. Elsir, M. F. E. R. O. Mohamed, A. B. Mahmoud, A. I. I. Ahmad, M. A. Adam, S. Altalib, O. A. DafaAllah, S. A. Hmed, A. S. Azman, I. Ciglenecki, E. Gignoux, A. González, C. Mwongera, M. A. Miranda, SARS-CoV-2 Antibody Prevalence and Population-Based Death Rates, Greater Omdurman, Sudan. Emerg. Infect. Dis. 28, 1026–1030 (2022).

19. EPHI PHEOC COVID-19. Ethiopian Public Health Institute, (available at https://ephi.gov.et/download/ephi-pheoc-covid-19/).

20. Official account of Yemen Supreme National Emergency Committee for COVID-19. Twitter, (available at https://twitter.com/ysneccovid19).

21. A. S. Iyer, F. K. Jones, A. Nodoushani, M. Kelly, M. Becker, D. Slater, R. Mills, E. Teng, M. Kamruzzaman, W. F. Garcia-Beltran, M. Astudillo, D. Yang, T. E. Miller, E. Oliver, S. Fischinger, C. Atyeo, A. J. Iafrate, S. B. Calderwood, S. A. Lauer, J. Yu, Z. Li, J. Feldman, B. M. Hauser, T. M. Caradonna, J. A. Branda, S. E. Turbett, R. C. LaRocque, G. Mellon, D. H. Barouch, A. G. Schmidt, A. S. Azman, G. Alter, E. T. Ryan, J. B. Harris, R. C. Charles, Persistence and decay of human antibody responses to the receptor binding domain of SARS-CoV-2 spike protein in COVID-19 patients. Sci Immunol. 5 (2020), doi:10.1126/sciimmunol.abe0367.

22. N. F. Brazeau, R. Verity, S. Jenks, H. Fu, C. Whittaker, P. Winskill, I. Dorigatti, P. G. T. Walker, S. Riley, R. P. Schnekenberg, H. Hoeltgebaum, T. A. Mellan, S. Mishra, H. J. T. Unwin, O. J. Watson, Z. M. Cucunubá, M. Baguelin, L. Whittles, S. Bhatt, A. C. Ghani, N. M. Ferguson, L. C. Okell, Estimating the COVID-19 infection fatality ratio accounting for seroreversion using statistical modelling. Commun. Med. 2, 54 (2022).

23. Yemen cemetery struggles to dig enough graves as coronavirus spreads. Reuters (2020), (available at https://www.reuters.com/article/uk-health-coronavirus-yemen-death-idUKKBN24A0XK).

24. UN Office for the Coordination of Humanitarian Affairs, Impact of COVID-19 on continuity of health services. Situation Reports (2020), (available at https://reports.unocha.org/en/country/sudan/card/49cURoYa8t/).

25. Sudan FETP Conducts Targeted Testing for COVID-19 in Khartoum State, (available at https://www.tephinet.org/sudan-fetp-conducts-targeted-testing-for-covid-19-in-khartoum-state).

26. O. Watson, N. Abdelmagid, A. Ahmed, A. E. Ahmed Abd Elhameed, C. Whittaker, N. Brazeau, A. Hamlet, P. Walker, J. Hay, A. Ghani, Others, Report 39: Characterising COVID-19 epidemic dynamics and mortality under-ascertainment in Khartoum, Sudan (available at https://spiral.imperial.ac.uk/bitstream/10044/1/84283/2/2020-12-01-COVID19-Report-39.pdf).

27. E. K. Gudina, S. Ali, E. Girma, A. Gize, B. Tegene, G. B. Hundie, W. T. Sime, R. Ambachew, Gebreyohanns, M. Bekele, A. Bakuli, K. Elsbernd, S. Merkt, L. Contento, M. Hoelscher, J. Hasenauer, A. Wieser, A. Kroidl, Seroepidemiology and model-based prediction of SARS-CoV-2 in Ethiopia: longitudinal cohort study among front-line hospital workers and communities. Lancet Glob Health. 9, e1517–e1527 (2021).

28. L. H. Nguyen, D. A. Drew, M. S. Graham, A. D. Joshi, C.-G. Guo, W. Ma, R. S. Mehta, E. T. Warner, D. R. Sikavi, C.-H. Lo, S. Kwon, M. Song, L. A. Mucci, M. J. Stampfer, W. C. Willett, H. Eliassen, J. E. Hart, J. E. Chavarro, J. W. Rich-Edwards, R. Davies, J. Capdevila, K. A. Lee, M. N. Lochlainn, T. Varsavsky, C. H. Sudre, M. J. Cardoso, J. Wolf, T. D. Spector, S. Ourselin, C. J. Steves, A. T. Chan, COronavirus Pandemic Epidemiology Consortium, Risk of COVID-19 among front-line health-care workers and the general community: a prospective cohort study. Lancet Public Health. 5, e475–e483 (2020).

29. R. Malaeb, N. Yousef, O. Al-Nagdah, Q. H. Ali, M. A. S. Saeed, A. Haider, E. Zelikova, N. Malou, S. Guiramand, C. Mills, F. Luquero, K. Porten, High Seroprevalence of Antibodies Against SARS-CoV-2 Among Healthcare Workers 8 Months After the First Wave in Aden, Yemen (2021),, doi:10.2139/ssrn.3883213.

30. R. K. Arora, A. Joseph, J. Van Wyk, S. Rocco, A. Atmaja, E. May, T. Yan, N. Bobrovitz, J. Chevrier, M. P. Cheng, T. Williamson, D. L. Buckeridge, SeroTracker: a global SARS-CoV-2 seroprevalence dashboard. Lancet Infect. Dis. 21, e75–e76 (2021).

31. Z. Munn, S. Moola, K. Lisy, D. Riitano, C. Tufanaru, Methodological guidance for systematic reviews of observational epidemiological studies reporting prevalence and cumulative incidence data. Int. J. Evid. Based Healthc. 13, 147–153 (2015).

32. A. Nkuba Ndaye, A. Hoxha, J. Madinga, J. Mariën, M. Peeters, F. H. Leendertz, S. Ahuka Mundeke, K. K. Ariën, J.-J. Muyembe Tanfumu, P. Mbala Kingebeni, V. Vanlerberghe, Challenges in interpreting SARS-CoV-2 serological results in African countries. Lancet Glob Health. 9, e588–e589 (2021).

33. United Nations General Assembly, Transforming our world: the 2030 Agenda for Sustainable Development. A/RES/70/1 (2015), (available at https://sdgs.un.org/2030agenda).

34. R. McCabe, O. J. Watson, mrc-ide/covid-alternative-mortality: v0.1.0 (2022; https://zenodo.org/record/7185171).

