## Supplementary Materials for "Alternative epidemic indicators for COVID-19: a model-based assessment of COVID-19 mortality ascertainment in three settings with incomplete death registration systems"

*For the purpose of open access, the author has applied a 'Creative Commons Attribution (CC BY) licence (where permitted by UKRI, 'Open Government Licence' or 'Creative Commons Attribution No-derivatives (CC-BY-ND) licence' may be stated instead) to any Author Accepted Manuscript version arising.*

#### Materials and Methods

We explain the general methodology, applicable to each of the three settings, before expanding on the specific data sets and sources for each modelled city. A schematic diagram of the methods is shown in Figure S1. All data and code are provided at https://github.com/mrc-ide/covid-alternative-mortality/ (*1*).

##### Mathematical model of SARS-CoV-2 transmission and disease progression

We use a previously published age-structured SARS-CoV-2 transmission model (*2*) and fitting framework (*3*) to fit the weekly estimated excess deaths in each setting in our study. In overview, the model is a Susceptible-Exposed-Infected-Recovered-Susceptible compartmental model, which is population-based and age-structured. The model explicitly represents disease severity and resultant passage through different healthcare levels, with an assumed elevated severity when healthcare capacity is exceeded as defined in Walker et al. (*2*). The model is capable of modelling vaccinations (see (*4*)), but in all settings considered in this study, vaccination campaigns had not started.

Model fitting was carried out within a Bayesian framework using a Metropolis-Hastings Markov Chain Monte Carlo (MCMC) based sampling scheme, which estimates the epidemic start date, $R_0$, and the time varying reproduction number, $R_t$, using a series of pseudo-random walk parameters ($\rho_n$), which alter transmission every 2-weeks, given by:

$$R_t = R_0 . f(-\rho_1 - \rho_2 ... - \rho_n)$$

where $f(x) = 2 . exp(x) / (1 + exp(x))$. Each random walk parameter is introduced two weeks after the previous parameter, serving to capture changes in transmission every two weeks. The last change in transmission, $n$, is maintained for the last 4 weeks prior to the current day to reflect our inability to estimate the effect size of this parameter due to the approximate 21-day delay between infection and death.

Each model fit is tailored to each setting, incorporating demographics, with the population size in 5-year age bands, and the effective number of general hospital beds and intensive care beds for each city.

#### Estimation of seroprevalence

Seroprevalence over time was derived from the total number of infections estimated by the model, adjusted for rates of seroconversion, seroreversion and serological assay sensitivity.

In each setting, we assumed that seroconversion followed an exponential distribution with mean time to seroconversion of 13.3 days for IgG antibodies (*5*). Seroconversion for IgM antibodies was assumed to occur more quickly, with mean time to seroconversion of 12.3 days (*5*). Similarly, in each setting seroreversion was assumed to follow a Weibull distribution with shape parameter 3.7 (*6*) and with the scale parameter adjusted to enforce a specific half-life for IgG or IgM antibodies, respectively. As default, we assumed a half-life of 50 days and 140 days for IgM and IgG, respectively, as estimated in Brazeau et al. (*6*). In Addis Ababa, a negligible amount of time had passed between the start of the epidemic inferred from the alternative mortality source and the serosurvey period and consequently the assumed duration of seroreversion does not impact our findings. However, because in Aden there was a substantial amount of time between the excess mortality and seroprevalence studies, we instead conducted a sensitivity analysis by varying the half-life of IgG antibodies between 100 and 280 days and of IgM antibodies between 30 and 70 days in order to capture the uncertainty surrounding the true value of these parameters. Lastly, in Khartoum, the observed seroprevalence estimated by Moser et al. (*7*) for Omdurman has already been adjusted to account for decreasing diagnostic test sensitivity over time as a result of waning antibody titres, which resulted in an increase from 34.3% crude seroprevalence to an adjusted seroprevalence of 54.6% (*7*). Consequently, we estimate seroprevalence from our model using a different approach. We continue to model the lag from infection to seroconversion, with a mean of 13.3 days. However, we consider individuals to only become seronegative again once they have moved from the recovered infection compartment to the susceptible compartment (i.e. by definition they should not possess antibodies to detect), which has a mean duration of 365 days and is described by an Erlang-2 distribution, i.e. is the sum of two independent exponential distributions each with a mean duration of 365/2 days.

Seroreversion and seroconversion distributions for combined IgG and IgM antibodies was estimated by treating the IgG and IgM distributions as independent. Although this assumption is unlikely to hold in reality, we validated our approach using Monte Carlo simulation which produced an essentially identical probability distribution of seropositivity when the two antibody types were dependent as to that when they were treated independently (Figure S2).

#### Testing for differences between observed and expected seroprevalence

Chi-squared tests were used to determine whether or not the difference between observed and expected seroprevalence was statistically significant.

Let $\theta_1$ denote reported seroprevalence and let $\theta_2$ denote our modelled seroprevalence. Our test statistic is:

$$T = \frac{(\theta_1 - \theta_2)^2}{var(\theta_1) + var(\theta_2\ )},$$

Where $var(\theta_1) \approx (\frac{upper\ confidence\ limit - lower\ confidence\ limit}{2 \times 1.96})^2$ , with 95% confidence limits those reported in the serosurveys.

Under the null hypothesis $T \sim \chi_1^{\ 2}$ , where $\chi_1^{\ 2}$ denotes a Chi-squared distribution with 1 degree of freedom, and we assume that there are no statistically significant differences between $\theta_1$ and $\theta_2$. A p-value greater than $0.05$ indicates that there is not enough evidence to reject the null hypothesis, and we conclude that there are no statistically significant differences between the reported and modelled values.

#### Addis Ababa

##### COVID-19 deaths in Addis Ababa

At the time of writing, we were unable to obtain a fully complete daily time series of confirmed COVID-19 deaths in Addis Ababa. Instead, we estimated this using information published by the Ethiopian Public Health Institute and trends in national cases in Ethiopia reported to the World Health Organization (WHO) (*8*). Based on these data, we estimated daily reported COVID-19 deaths for three distinct time periods before combining into one time series for 2020.

The first confirmed death in Ethiopia was registered on 6 April 2020 and was reported as having occurred in Addis Ababa (*9*). By 21 June 2020, Watare et al. (*10*) reported a total of 63 COVID-19 deaths in Addis Ababa compared to 72 at a national level (87.5%). Therefore, the first time period considered was 6 April - 21 June 2020 and the daily time series was estimated by assuming that each day, the number of deaths in Addis Ababa was 87.5% of the deaths at a national-level rounded to the nearest integer. This produced 66 deaths in total for Addis Ababa for this period, and so three dates were randomly selected from this period and one death was removed from that date's death toll.

The Ethiopian Public Health Institute (EPHI) (*8*) reported a total of 295 cumulative COVID-19 deaths in Addis Ababa as of 9 August 2020, resulting in 232 deaths in Addis Ababa during this period compared to 318 nationally (73%). Using the same method as above, the second time period for which a daily time series was constructed was 22 June - 9 August 2020. In this instance, there were two fewer deaths than the total and two dates on which national deaths were recorded were randomly selected in which to add one additional death to.

Finally, from 10 August 2020 the EPHI began publishing weekly COVID-19 mortality figures for Addis Ababa (with the exception of the week commencing 19 October, for which the weekly total was assumed to be the median, rounded to the nearest integer, of the two surrounding weeks). The time period from 10 August until 31 October 2020 was the third period under consideration. Within each week, the daily number of COVID-19 deaths was assumed to follow the same proportion of deaths falling on that weekday at national level. The three time series were then joined into a coherent time series from 6 April - 31 October 2020.

##### Excess deaths in Addis Ababa

Excess mortality was estimated using cemetery surveillance data gathered by Endris et al. (*11*), which detailed the number of burials per day at all cemeteries in Addis Ababa across January

2015 - January 2021. We considered two mortality baselines using data from 2015 - 2019 and exclusively from 2019, respectively. We fit a negative binomial distribution to the total number of daily burials across cemeteries per calendar month. Baseline mortality was then estimated by sampling 100 times from each distribution per date, with excess mortality calculated per sample by taking the difference between the observed and baseline number of deaths per day. Our finalised time series of excess mortality using these data was averaged over each sample and presented as the rolling 7-day mean.

#### Seroprevalence in Addis Ababa

We used estimates of seroprevalence in Addis Ababa from Abdella et al. (*12*), who conducted a serosurvey of 956 individuals to detect both IgG and combined IgG/IgM antibodies in this area across 22 July - 10 August 2020 using the *Core Technology IgM/IgG rapid test*. This assay has a manufacturer reported sensitivity of 91.4% and 89.4% for IgG and IgM antibodies, respectively. Although confidence intervals are not reported by the manufacturer, Abdella et al. (*12*) report the results of an independent trial of 200 samples in which IgG and IgM specificity was estimated as 97% (95% CI 94.6% - 99.4%) and 98.0% (95% CI 96.0% - 99.9%), respectively. For the purposes of our modelling we used the sensitivity and specificity values estimated by the manufacturer.

We were unable to obtain specific seroconversion and seroreversion rates for the assay used in Abdella et al.'s (*12*) study and therefore assumed our default parameters as set out above. Although we were unable to explain the large difference observed between IgG and combined IgG/IgM antibodies, even when varying the seroreversion half-life of IgM, we hypothesise that this could be attributed to lack of validation of the assay in low-income countries.

#### Demography and healthcare in Addis Ababa

Age-stratified estimates of the population of Addis Ababa were derived by assuming that the proportions of individuals in each age group were those observed by Timotewas et al. (*13*) in 2012-2013 (summed across male and females), and multiplied by the most recent estimate of the population of Addis Ababa in 2020 (4.8 million) (*14*).

Laytin et al. (*15*) estimated 130 intensive care unit (ICU) beds in Addis Ababa in January 2020. Assuming that ICU beds are 1.5% of all hospital beds in low-income countries (*16*), we estimated there to be 6867 acute hospital beds in Addis Ababa.

Model fits and estimated seroprevalence

We fit the model up to 31 October 2020, by which point the serosurvey had taken place and the first wave of reported COVID-19 deaths had plateaued. Figures S4 - S6 present the model fits to COVID-19 and excess deaths under different baselines and the associated model-estimated seroprevalence.

Figure S7 is analogous to Figures 1C and 1D in the main text, but also includes the unweighted seroprevalence reported by Abdella et al. (*12*). The unweighted seroprevalence estimates are greater than their corresponding weighted estimates. The unweighted estimates do not account for the demography of the sampled population. When comparing our modelled seroprevalence to the reported unweighted seroprevalence, there are fewer scenarios that are not statistically different when compared to the weighted seroprevalence. Scenarios that were not statistically different included mortality derived using data from all years to inform the baseline and using 2019 from June 2020 onwards produced estimates of IgG and combined IgG/IgM antibodies consistent with the unweighted results in Abdella et al. (*12*) (p-values: 0.779; 0.487; 0.282; 0.925, respectively), as did the model fit to reported COVID-19 deaths for IgG antibodies only (p-value: 0.112).

#### Aden

##### COVID-19 deaths in Aden

At the time of writing, we were also unable to obtain from a formal ministry of health source the daily time series of confirmed COVID-19 deaths in Aden. Instead, we scraped the cumulative number of deaths from tweets directly referencing Aden from the Twitter account of the Yemen Supreme National Emergency Committee for COVID-19 (*17*) and converted this into a daily time series for reported COVID-19 deaths.

##### Excess deaths in Aden

We used excess mortality in Aden as estimated by Koum Besson et al. (*18*), who derived estimates of excess mortality using satellite images of all active cemeteries in Aden. We converted the cumulative excess data into a daily time series and applied a rolling 7-day mean to the daily estimated number of excess deaths.

##### Seroprevalence in Aden

Estimates of the seroprevalence of combined IgG/IgM antibodies in Aden across 28 November - 13 December 2020 were sourced from Bin-Ghouth et al. (*19*), using the *Healgen COVID 19 IgG/IgM Rapid Test Cassette* and confirmed via the WANTAI SARS-CoV-2 Ab ELISA (China). They recruited 2,001 participants to their study. The reported ELISA sensitivity and specificity by the manufacturer are: IgG sensitivity = 96.7% (83.3% - 99.4%), IgM sensitivity = 100% (88.7% - 100%). In the absence of assay-specific estimates of rates of seroconversion, we used the default assumptions as set out above. However, as noted in the main text, due to the long delay between the peak in excess mortality and the implementation of the serosurvey in addition to the lack of assay-specific estimates of rates of seroreversion, we conducted a sensitivity analysis of different IgG and IgM seroreversion half-lives to assess the uncertainty in these parameters.

##### Demography and healthcare in Aden

We used an analogous methodology to derive age-stratified estimates of the population of Aden as detailed above for Addis Ababa. We used the population pyramid provided by Bawazir (*20*). We split the age category ‘75+’ (in years) into ‘75 - 79’ and ‘80 - 84’, as required by our model, by assuming the proportions of individuals within these two categories followed that estimated nationally by the United Nations World Population Prospects 2019 (*21*). We multiplied this by the estimated population of Aden (1 million) as assumed in Koum-Besson et al. (*18*).

Official data on healthcare capacity in Aden were not available at the time of writing. International media reported 18 ICU beds in Aden in June 2020 (*22*). We verified this figure by assuming that the reported 520 ICU beds in Yemen were spread across the country proportional to the population of each area. Assuming Aden accounts for 3.35% of the population (total population of Yemen 29.83 million (*21*)), this implies 17 ICU beds in this area. Under the same assumption of 1.5% of hospital beds are ICU beds as above in Murthy et al. (*16*), we estimate there to be 1133 acute hospital beds in Aden.

#### Model fits and estimated seroprevalence

Figure S8 presents the model fit to reported COVID-19 deaths and the resulting estimated seroprevalence of IgM, combined IgG/IgM and IgM antibodies over time. We present estimated seroprevalence under the minimum and maximum seroreversion IgG and IgM half-lives included in our sensitivity analysis, 100 days and 280 days for IgG and 30 days and 70 days for IgM, neither of which can explain the observed seroprevalence.

As our excess mortality time series ended on 16 September 2020 (the end of Koum Bessom et al.'s study (*18*)), we used our model to project forward the number of daily deaths from this point to the end of 2020 assuming that $R_t$ remained at the level observed on this date. This is illustrated in Figure S9. Figure S10 presents multiple model fits to the Aden epidemic under different assumptions of the population average IFR. These figures indicate that an IFR of 0.2% is unable to recreate the wave of excess mortality in Aden, which is due to the depletion of the susceptible population.

Figure S11 shows the results of a sensitivity analysis of varying the IgM half-life between 30 and 70 days, of which there is minimal impact on the results in comparison to the reported values for IgM and combined IgG/IgM antibodies.

#### Khartoum

##### COVID-19 deaths in Khartoum

COVID-19 deaths for Khartoum were scraped from daily epidemiological reports published by the Sudan Health Observatory (*23*). On 8 November 2020, the Sudanese Federal Ministry of Health (FMOH) released a periodic revision of COVID-19 data, which included an additional 99 deaths for Khartoum, increasing the cumulative total COVID-19 deaths to 414 by 8 November. These additional 99 deaths were reported to have occurred any time since the beginning of the pandemic. In response, we distributed the additional 99 deaths in proportion to the weekly mean number of COVID-19 deaths reported in the daily epidemiological reports, which ensures the same shape of the epidemic but with an increased magnitude. Lastly, on 17 November 2020, the FMOH released a statement detailing that an additional 46 deaths had been reported from private diagnostic laboratories and had occurred between 8 November 2020 and 17 November 2020 (*24*). During this period, only 1 death had been reported in FMOH epidemiological reports, however, it was subsequently followed by a sharp, exponential increase in reported COVID-19 deaths in epidemiological reports. In response, we assumed that the additional 46 deaths were exponentially increasing between 8 November and 17 November 2020, which also aligns with the increase in reported COVID-19 cases in epidemiological reports.

##### Symptomatic cases  in Khartoum

We used estimates of the cumulative number of symptomatic COVID-19 cases in individuals over the age of 15 in Khartoum, which were estimated using an online survey as described in Mohamed et al. (*25*). The survey form was distributed through social media channels in Khartoum between 26 May to 3 June 2020 and collected information on whether participants had suffered symptoms associated with COVID-19 since the start of the pandemic. Symptomatic individuals were subsequently asked to report which of any of 13 different symptoms they had suffered and all individuals were asked both i) if they had received a SARS-CoV-2 diagnostic test and ii) the outcome of any test taken. From this survey, the authors created a statistical model to infer the SARS-CoV-2 infection status of survey participants who had not received a SARS-CoV-2 test based on reported symptoms and through this, infer the symptomatic attack rate in the general population by 3 June. We use three estimates of the symptomatic attack rate in 15+ year olds from the study to reflect the uncertainty in the inferred attack rates: 8.3%, 11.0% and 13.7%.

##### Seroprevalence and mortality survey in Khartoum

We leveraged a cross-sectional, household survey in Omdurman, Sudan, conducted during March-April 2021 by Médecins sans frontières (*7*). Omdurman is the largest city in terms of population within Khartoum state and represents approximately one-third of the population of Khartoum state. All members of a subset of households, regardless of age were invited to participate in a seroprevalence survey, which recruited 3808 individuals. The survey used the SD-Biosensor STANDARD Q COVID-19 IgM/IgG Combo rapid diagnostic test (RDT), with a subset of dried blood spots also collected for analysis by ELISA (Anti–SARS-CoV-2 ELISA [IgG, S1 domain]; Euroimmun). In the analysis conducted by Médecins sans frontières (*7*), crude seroprevalence estimates from the RDT and ELISA results were adjusted using published performance estimates for the RDT and ELISA in a meta-analysis with random effects model before being used as inputs for a Bayesian latent class model to yield adjusted seroprevalence estimate of 54.6% (95% CI 51.4–57.8).

In the same household survey, a mortality survey was conducted to investigate the death rate for the pre–pandemic (January 1, 2019–February 29, 2020) and pandemic (March 1, 2020 – day of the survey). Data from 27,315 people across 3716 households collected across the entire recall period showed a 67% (95% CI: 32–110) increase in crude death rate between the pre–pandemic (0.12 deaths/10000 people/day [95% CI: 0.10 – 0.14]) and pandemic (0.20 [95% CI: 0.16 – 0.23]) periods. This equates to a 7113 excess deaths across Omdurman (95% CI: 5015 – 9505). To compare this against the predicted COVID-19 deaths from our transmission models fit to data across all Khatoum state, we linearly scale these estimates by 292% to reflect the larger population size of Khartoum state (estimated to be 8,877,146 by the United Nations Office for the Coordination of Humanitarian Affairs) compared to Omdurman (*7*).

##### Demography and healthcare in Khartoum

We assume the total population size for Khartoum state is 8877146, as estimated by UN OCHA (*26*), with a demographic profile comparable to Sudan as estimated in the United Nations World Population Prospects 2019 (*21*). We assume there are 4640 general hospital beds based on recent per capita estimation by the non-profit organisation NextGen Sudan in October 2019 (*27*), and that 75 ICU beds are available based on Sulieman et al. (*28*).

#### Estimation of reporting fractions

Reporting fractions are defined as total COVID-19 deaths over the period divided by total positive excess deaths over the same period (Table S5).

#### Comparison of reported and WHO-estimated excess mortality

Using excess mortality estimated by the WHO, we were able to compare reporting fractions of COVID-19 deaths under the alternative sources used in each setting to those at a national level (Figure S15). These data suggest 8.8% (95% CI: 4.3% - 26.1%), 5.6% (95% CI: 3.5% - 9.3%) and 10.0% (95% CI: 6.4% - 16.4%) of COVID-19 deaths were captured within official statistics in Ethiopia, Yemen and Sudan, respectively (Table S6). These estimates most likely should not align with our estimated reported fractions, with differences between local and national-level estimates to be expected due to regional heterogeneity, for example, in access to healthcare and death registration services in rural versus urban settings. However, we are encouraged to see similar magnitudes being estimated by our analyses in Aden (parameter uncertainty range: 0.8% - 8.0%) and Khartoum (parameter uncertainty range: 3.0% - 6.0%), which are slightly lower than suggested by the corresponding national-level figures. However, we estimate a much greater reporting fraction in Addis Ababa (parameter uncertainty range: 68.7% - 100%) than the corresponding national estimate, suggesting that national-level excess mortality may have been overestimated if national patterns are comparable to those in Addis Ababa. One plausible explanation for this relates to the WHO methodology for estimating excess mortality. These estimates are modelled based on pooling information from countries with similar characteristics, including WHO region, income status and burden of HIV (*29*). However, only four African Region (AMRO) and four Eastern Mediterranean Region (EMRO) countries could provide complete national all-cause mortality data. Consequently, WHO reports very wide confidence intervals for their estimates of excess mortality (Table S6). These wide ranges demonstrate the value in the alternative data sources we have used here to provide a more nuanced perspective of COVID-19 mortality reporting in low-income settings and add additional data that can be used to refine future estimates of excess mortality.

#### Supplementary Figures

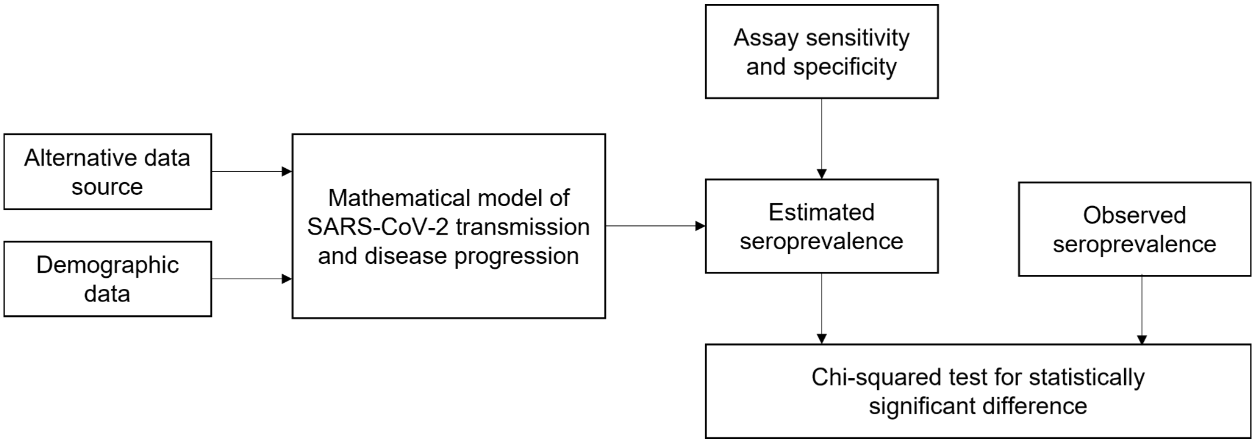


**Figure S1: Schematic diagram of the methodology used in each setting.** Within each setting, the SARS-CoV-2 transmission model used in this analysis is calibrated using the alternative data source and demographic data. From the model and assay-specific sensitivity and specificity, we estimate seroprevalence and compare this to that which was observed in independent studies in each setting. Finally, a Chi-squared test is used to test whether the difference between estimated and observed seroprevalence is statistically significant.

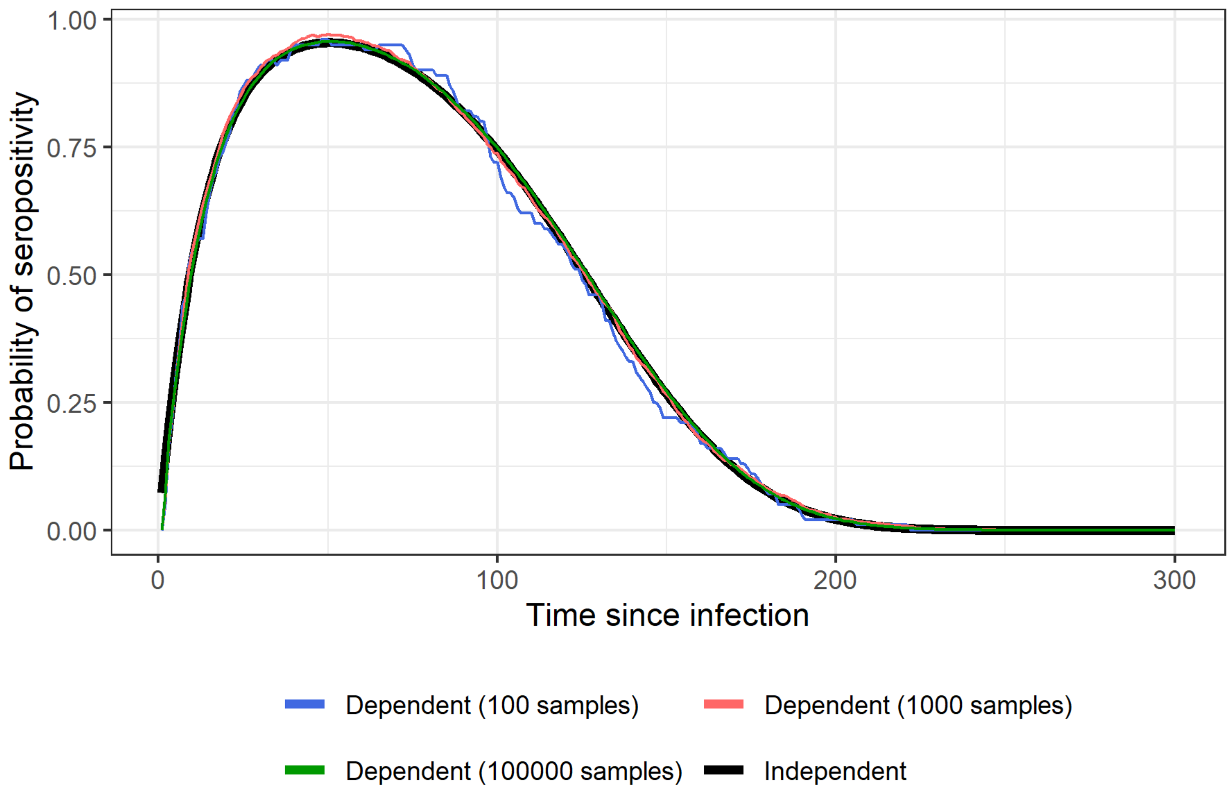


**Figure S2:** Probability of seropositivity with and without the assumption of independence between IgG and IgM antibodies.

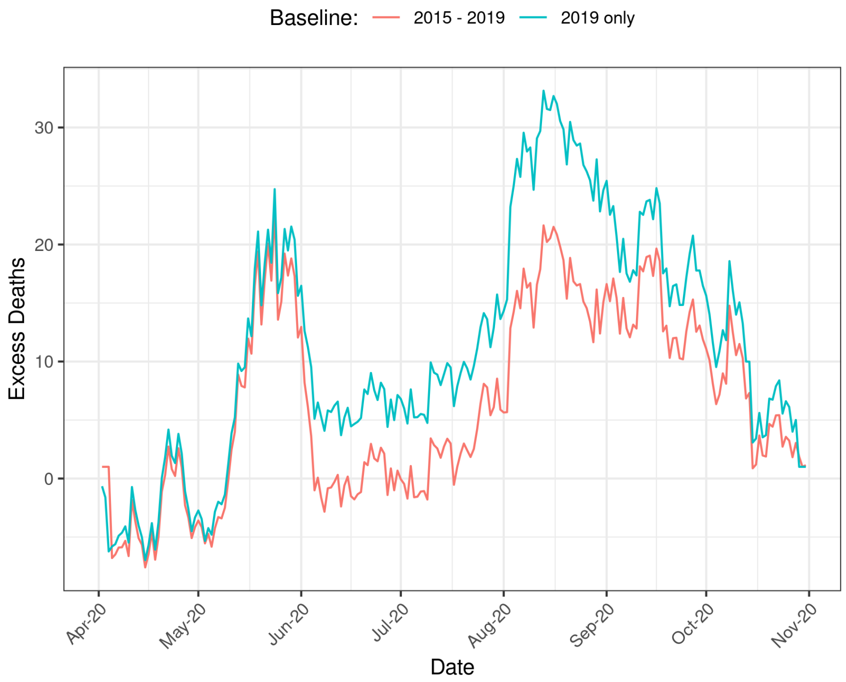


**Figure S3: Inferred excess deaths in Addis Ababa.** Excess deaths between April 2020 and November 2020 are shown for two methods (using all years between 2015-2019, or just 2019) of calculating the assumed baseline mortality during 2020 in the absence of the pandemic.

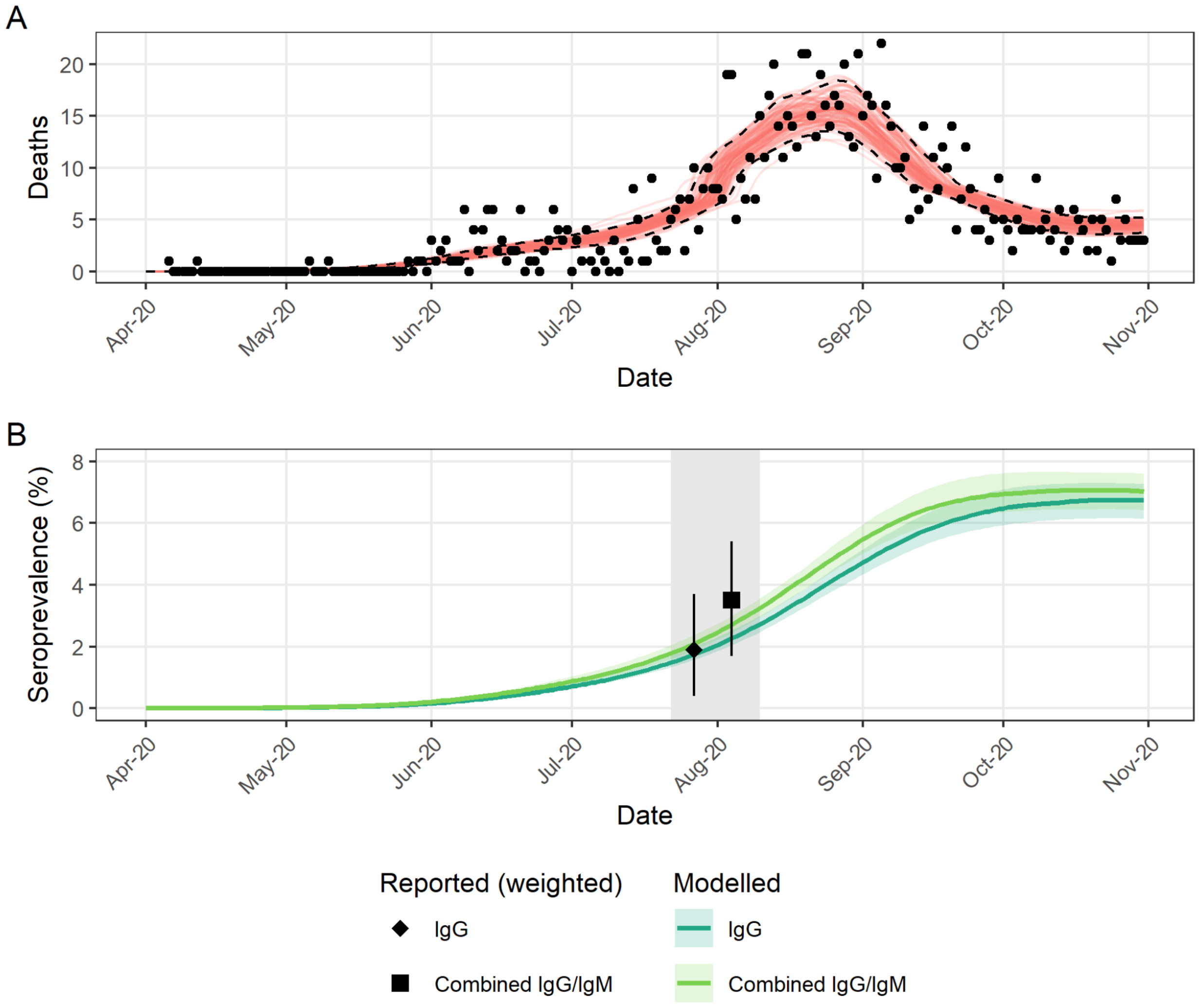


**Figure S4:** (A) Model fit (red lines) to reported COVID-19 deaths (black points) in Addis Ababa. (B) Estimated seroprevalence from model fit to COVID-19 deaths in (A) (median and 95% credible intervals) compared to reported values by Abdella et al. (*12*). Grey shaded area highlights the sampling period of the serosurvey, with weighted reported estimates both corresponding to this entire period.

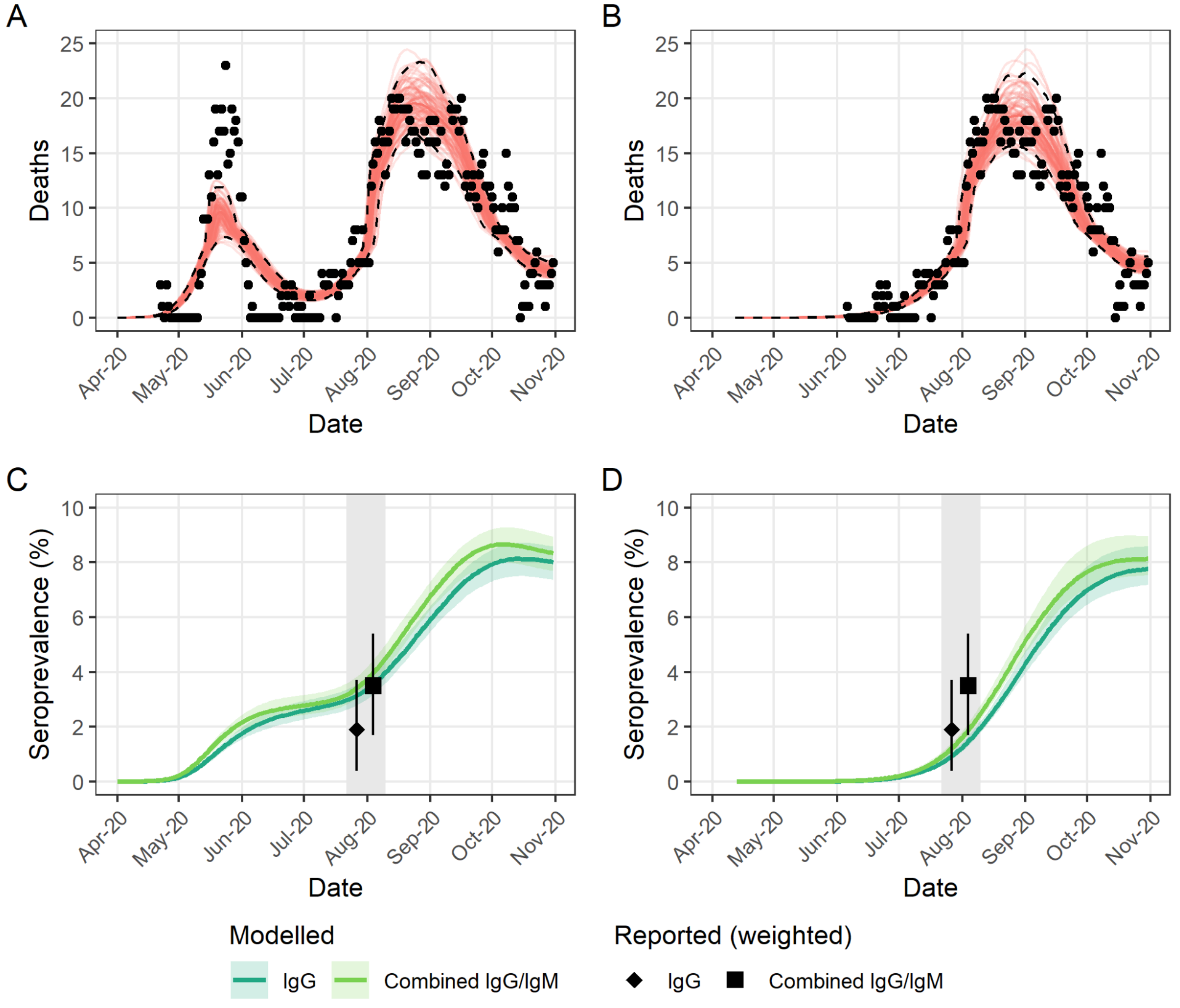


**Figure S5: Model fit and estimated seroprevalence under excess mortality estimated using data from 2015 - 2019 in Addis Ababa.** (A) Model fit to excess mortality. (B) Model fit to excess mortality without the first peak observed across May and June 2020. In (A) and (B), red lines represent model realisations and black dots show estimated excess deaths. (C) Estimated seroprevalence (median and 95% credible intervals) of IgG and combined IgG/IgM antibodies under model presented in (A). (D) Estimated seroprevalence (median and 95% credible intervals) of IgG and combined IgG/IgM antibodies under model presented in (B). In (C) and (D), black points indicate weighted seroprevalence as reported by Abdella et al. (*12*) and grey shaded area highlights the sampling period of the serosurvey, with weighted reported estimates both corresponding to this entire period.

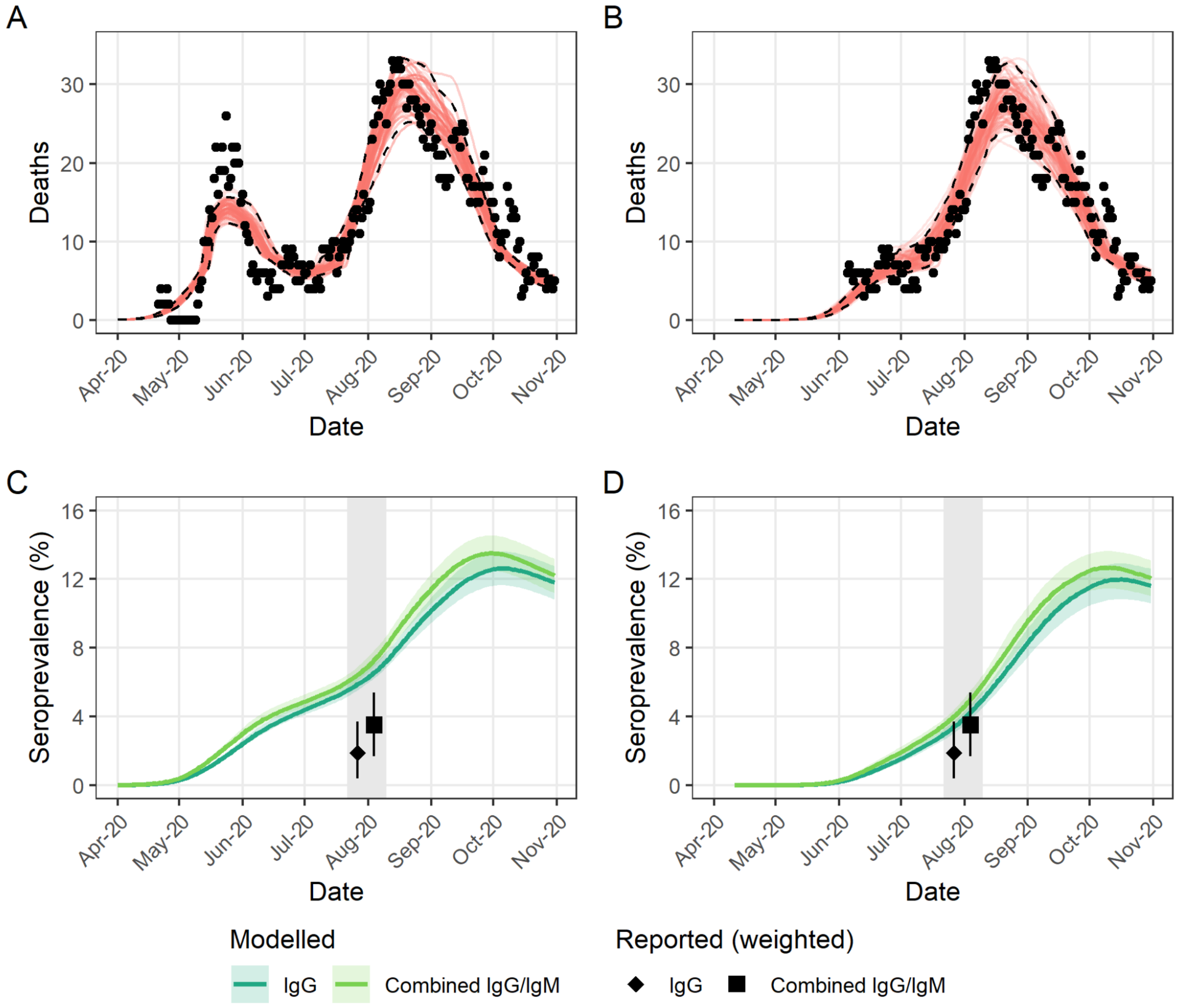


**Figure S6: Model fit and estimated seroprevalence under excess mortality estimated using data from 2019 only in Addis Ababa.** (A) Model fit to excess mortality. (B) Model fit to excess mortality without the first peak observed across May and June 2020. In (A) and (B), red lines represent model realisations and black dots show estimated excess deaths. (C) Estimated seroprevalence (median and 95% credible intervals) of IgG and combined IgG/IgM antibodies under model presented in (A). (D) Estimated seroprevalence (median and 95% credible intervals) of IgG and combined IgG/IgM antibodies under model presented in (B). In (C) and (D), black points indicate weighted seroprevalence as reported by Abdella et al. (*12*) and grey shaded area highlights the sampling period of the serosurvey, with weighted reported estimates both corresponding to this entire period.

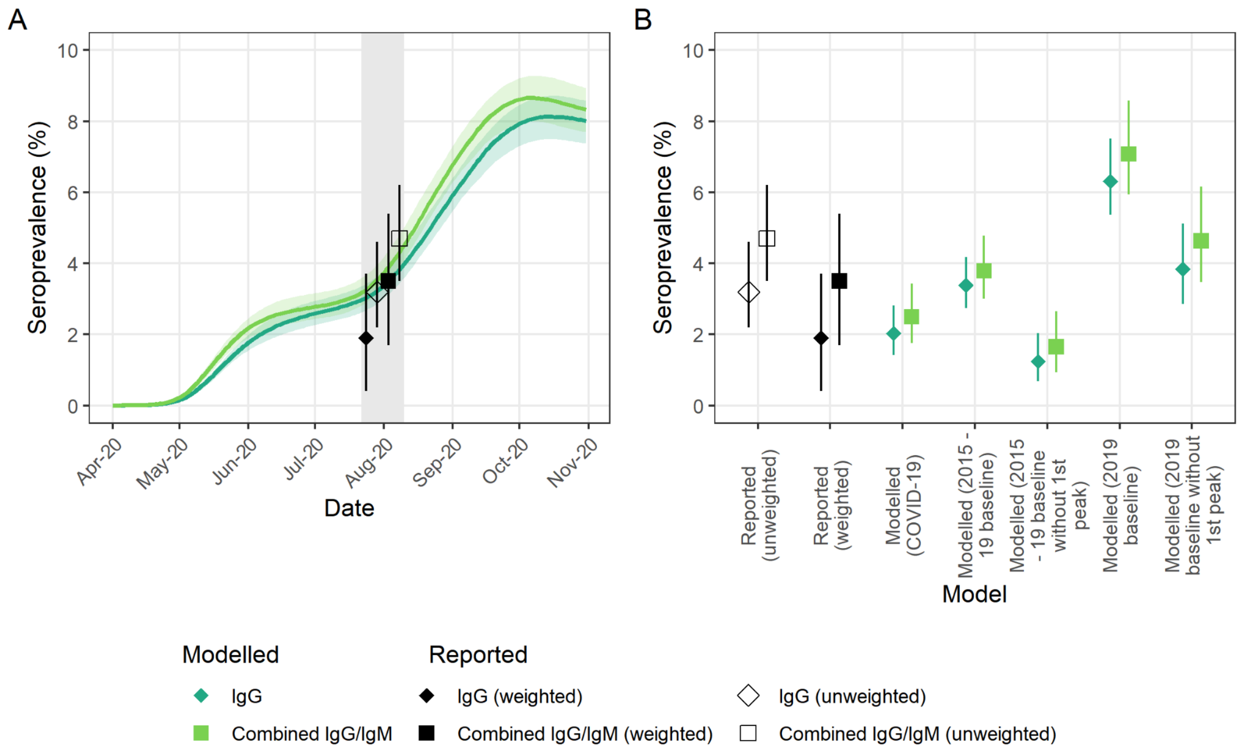


**Figure S7: Seroprevalence in Addis Ababa, Ethiopia in 2020 with reported weighted and unweighted values.** (A) Estimated seroprevalence from model fit to excess mortality under 2015 - 2019 baseline with the first peak compared to reported values from Abdella et al. (*12*). Grey shaded area highlights the sampling period of the serosurvey, with reported estimates both corresponding to this entire period. (B) Seroprevalence estimated under models fit to different estimates of mortality from COVID-19 and excess mortality with different baselines compared to reported seroprevalence from (*12*).

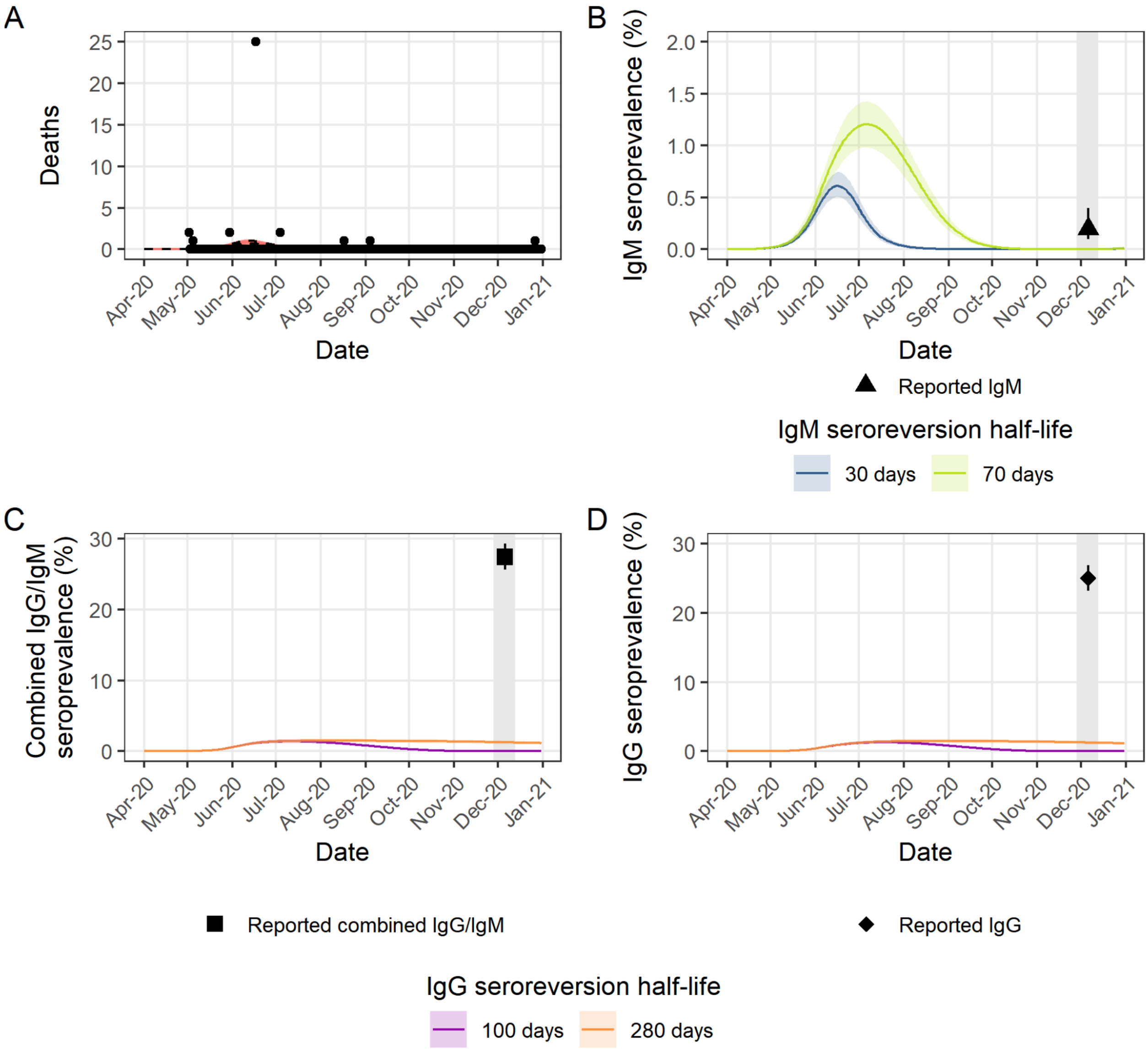


**Figure S8:** (A) Model fit to reported COVID-19 deaths in Aden under default infection fatality ratio (IFR=0.3%). Red lines indicate model realisations and black dots show reported COVID-19 deaths. (B) Estimated seroprevalence (median and 95% credible intervals) of IgM antibodies under minimum (30 days) and maximum (70 days) IgM sero-reversion half-lives included in our analysis compared to the observed value. Reported IgM by Bin-Gouth et al. (*19*) is 0.2% (95% CI: 0.1% - 0.4%). (C) Estimated seroprevalence (median and 95% credible intervals) of combined IgG/IgM antibodies under minimum (100 days) and maximum (280 days) seroreversion half-lives included in our sensitivity analysis compared to observed value. IgM half-life held constant at 50 days. Reported value is 27.4% (95% CI: 25.6% - 29.3%) (*19*). (D) As in (C) but for IgG antibodies. Reported value is 25.0% (95% CI: 23.2% - 26.9%) (*19*).

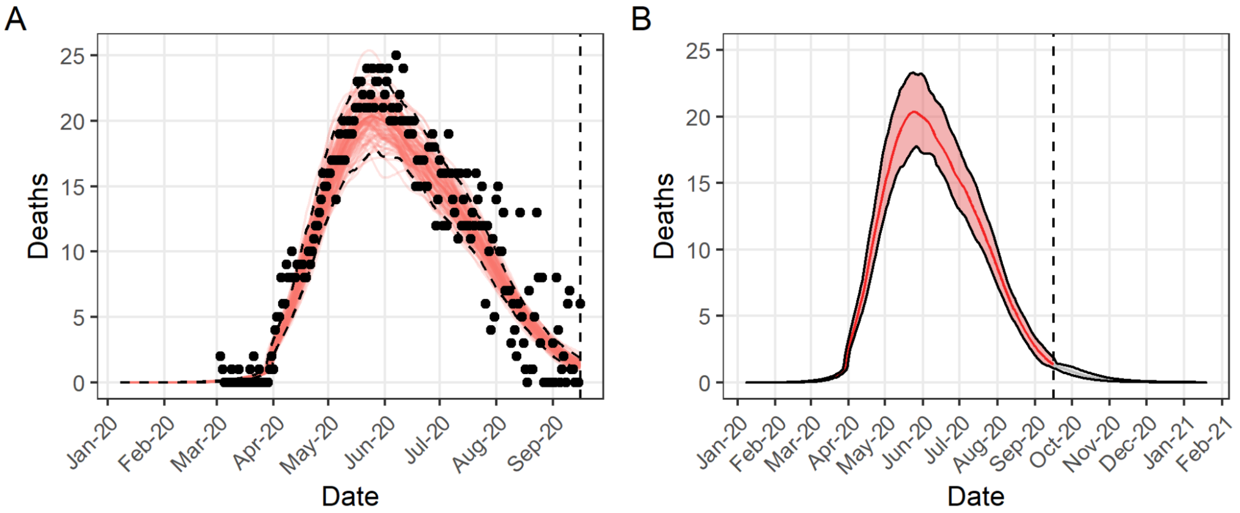


**Figure S9:** (A) Model fit to excess mortality in Aden. Red lines indicate model realisations and black dots show excess deaths. (B) Projection from mid-September (median and 95% credible intervals) to end of 2020 (grey) based on fit in (A) (red) assuming $R_t$ remains at value estimated on the last day of model fit (16 September 2020).

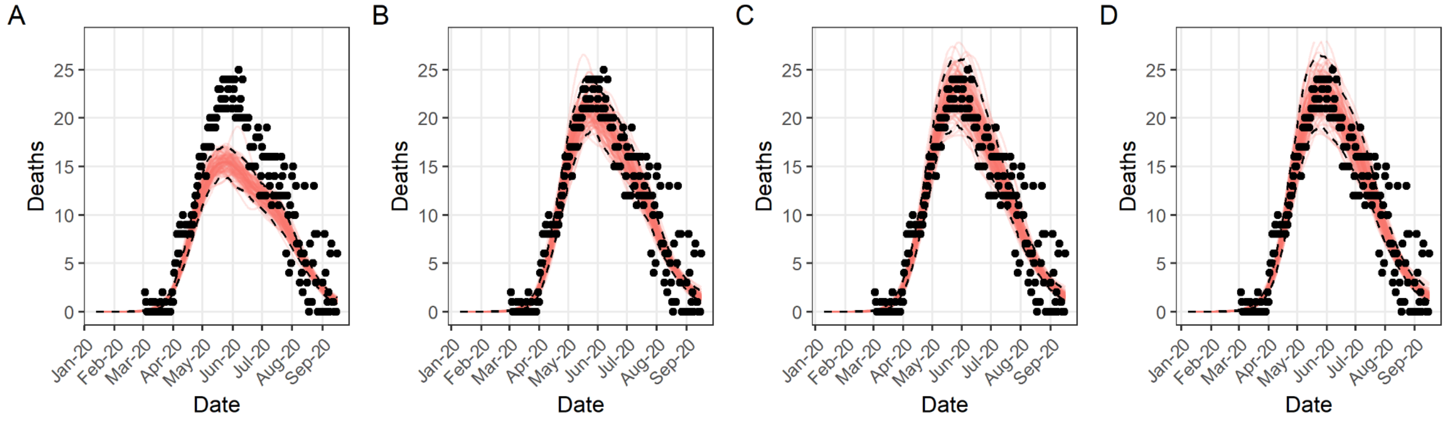


**Figure S10: Model fits to excess mortality in Aden under different assumptions of the Infection Fatality Ratio (IFR).** (A) IFR = 0.2; (B) IFR = 0.3; (C) IFR = 0.4; (D) IFR = 0.5. In all panels, red lines indicate model realisations and black dots show excess deaths.

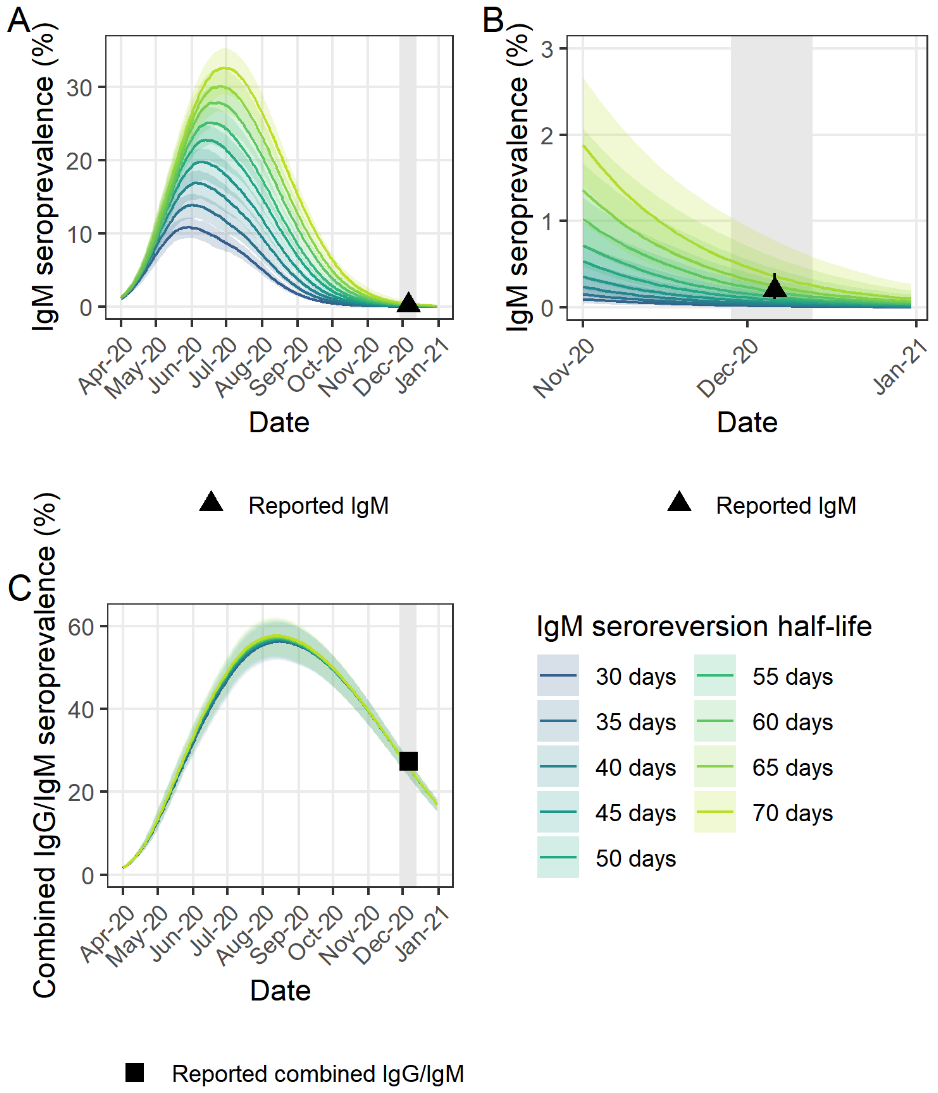


**Figure S11: Estimated seroprevalence in Aden under model fit to excess mortality under default infection fatality ratio (IFR=0.3%).** (A) Estimated seroprevalence of IgM antibodies in Aden under different assumptions of the IgM seroreversion half-life compared to that reported by Bin-Gouth et al. (*19*) (0.2% (95% CI 0.1% - 0.4%)). (B) As in (A) but restricted to November - December 2020 only. (C) Estimated seroprevalence of combined IgG/IgM antibodies under assumptions of the IgM seroreversion half-life compared to that reported in (*19*) (27.4% (95% CI 25.6% - 29.3%). IgG seroreversion half-life is held constant at 180 days.

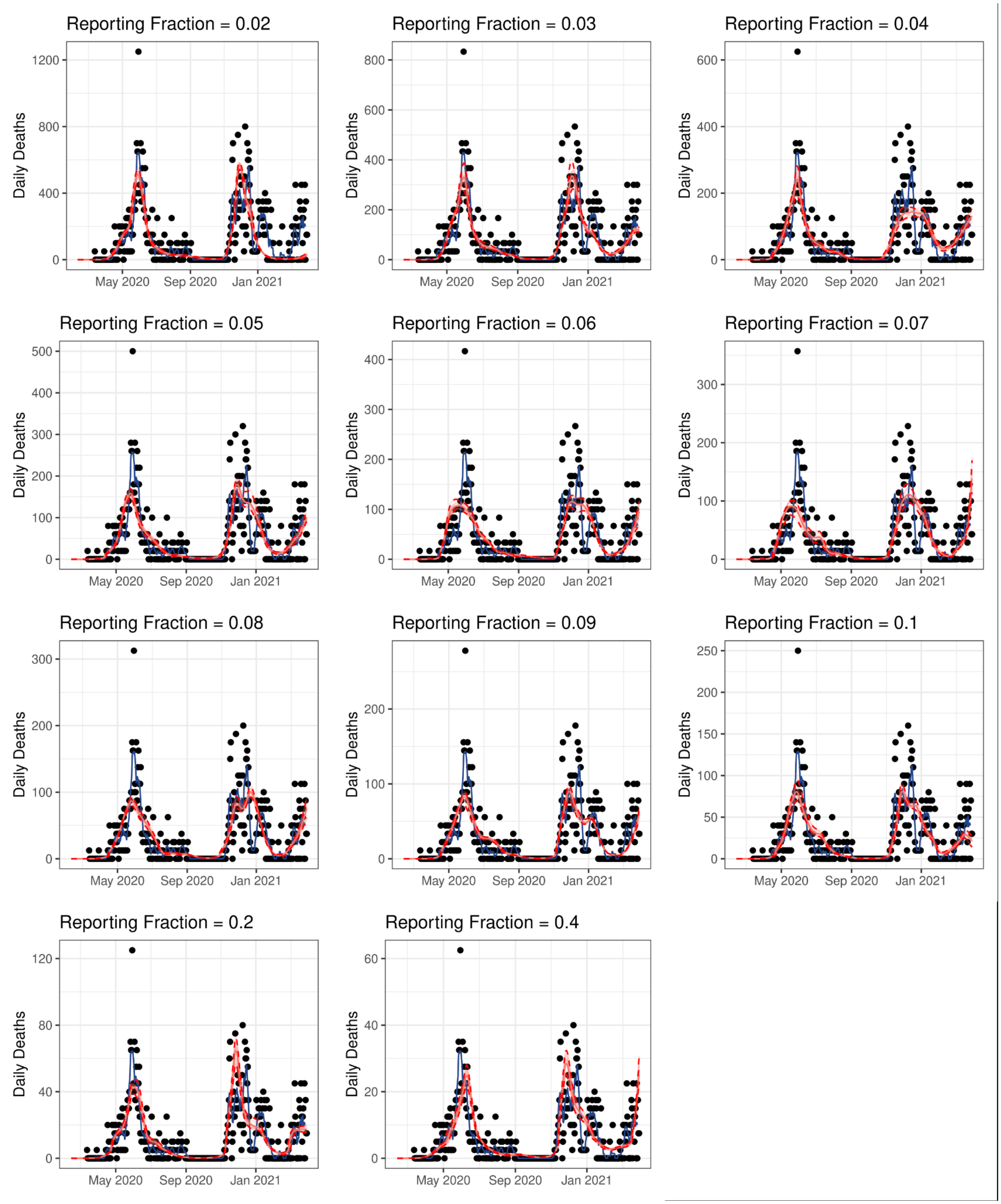


**Figure S12: Model fits in Khartoum based on different reporting fractions.** Black dots represent daily reported COVID-19 deaths, with the weekly mean shown with the blue line. Model fits are shown in red, with the 95% CI shown with red dashed lines.

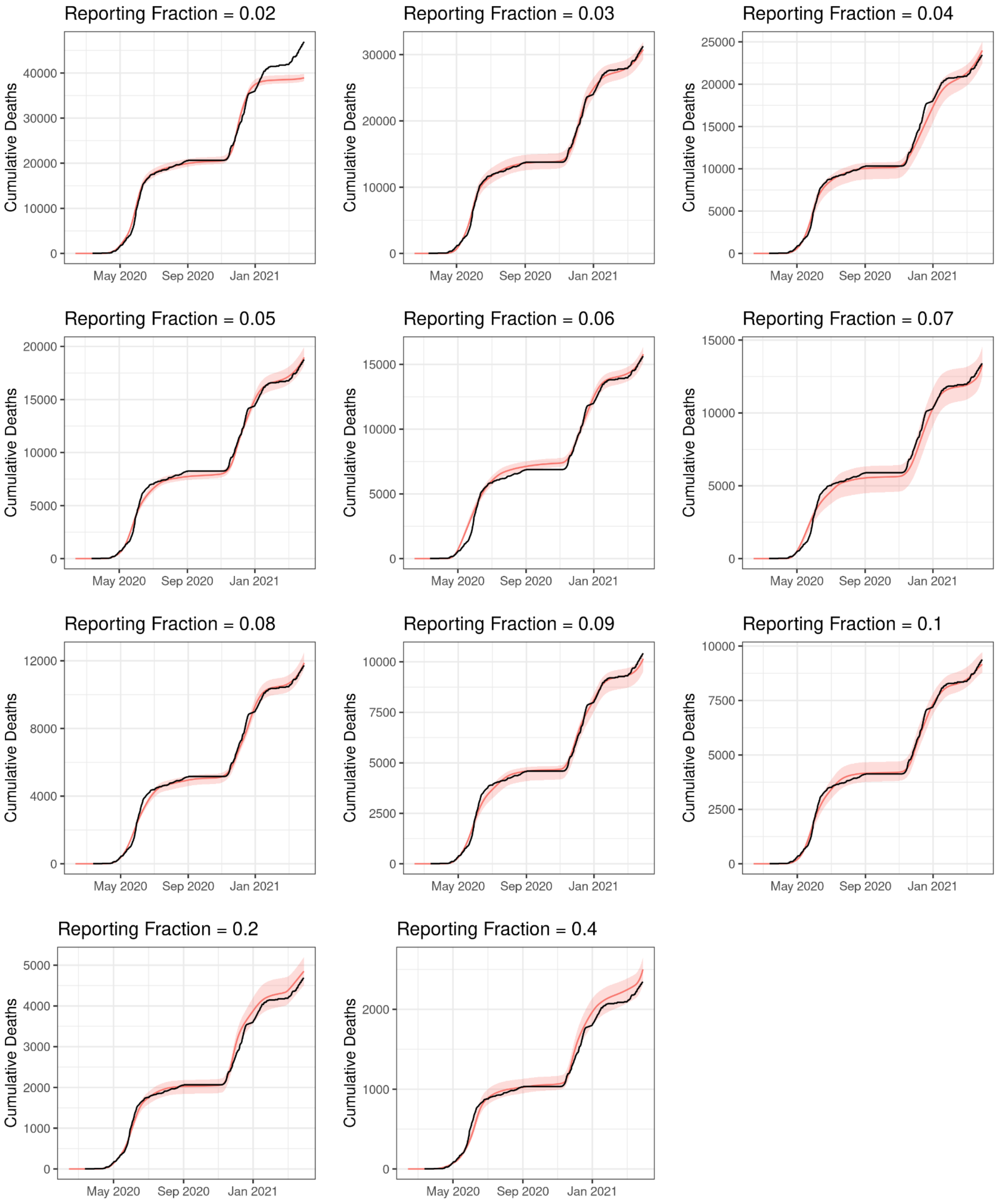


**Figure S13: Model fits in Khartoum based on different reporting fractions.** Black lines represent the cumulative reported COVID-19 deaths, with model fits shown in red (median and 95% CI).

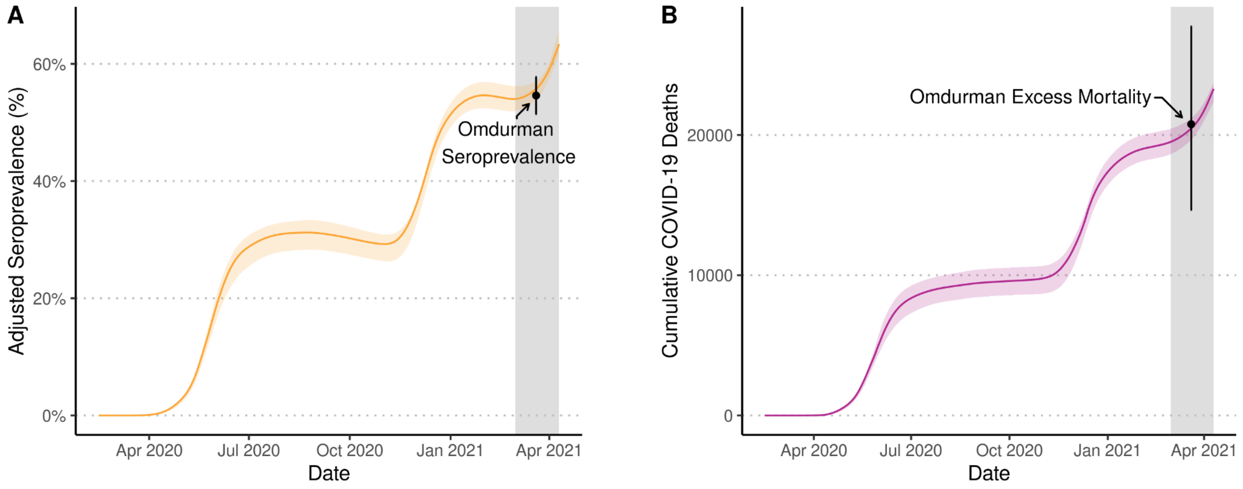


**Figure S14: Best model fit to Omdurman seroprevalence.** Model predicted **a)** adjusted seroprevalence and **b)** COVID-19 deaths are shown for Khartoum with a reporting fraction equal to 4.5%, which provided the best fit to the Omdurman data, which is shown in black (point estimates, 95% CI shown with vertical bars, and horizontal bars indicate date range for survey). The grey shaded area highlights the sampling period of the serosurvey and mortality survey in Omdurman.

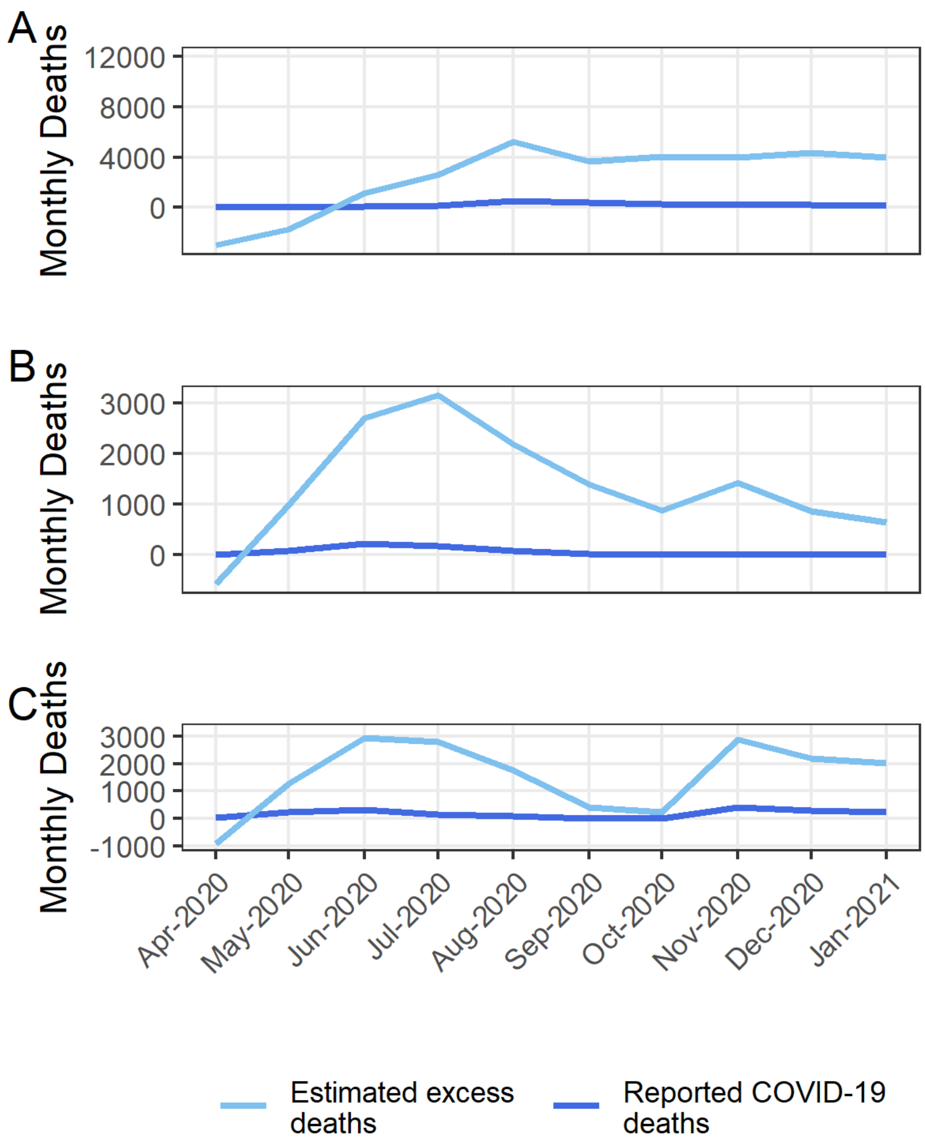


**Figure S15: WHO-estimated excess mortality** (*29*) **compared to reported COVID-19 deaths across April 2020 - December 2020.** (A) Ethiopia; (B) Sudan; (C) Yemen.

### Supplementary Tables

#### Table S1. Further details of alternative data collected at study locations and seroprevalence assays used.

| Setting | Alternative data source | | Seroprevalence | | | |
|---|---|---|---|---|---|---|
| | Overview | Ref | Overview | Assay details | Estimate | Ref |
| Addis Ababa, Ethiopia | Burial Site Worker Cemetery Reports.<br><br>01/01/2015 - 26/01/2021 | (*11*) | Random sample of 956 households. 22/07/2020 - 10/08/2020 | Core Technology IgM/IgG rapid test.*<br><br>IgG: sensitivity 91.4%; specificity 98.6%<br><br>IgM: sensitivity 89.4%; specificity 98.4% | IgG:<br>1.9% (0.4% - 3.7%)<br><br>Combined IgG/IgM:<br>3.5% (1.7% - 5.4%) | (*12*) |
| Aden, Yemen | Satellite Imagery of cemetery burials.<br><br>21/07/2016 - 19/09/2020 | (*18*) | Cross-sectional household study of 2001 people. 28/11/2020 - 13/12/2020 | Healgen COVID 19 IgG/IgM Rapid Test Cassette & confirmed via enzyme-linked immunosorbent assay (ELISA)<br><br>IgG: sensitivity 96.7% (83.3% - 99.4%)<br><br>IgM: sensitivity 100% (88.7% - 100%)<br><br>IgG or IgM: sensitivity 100% (88.7% - 100%); specificity 97.5% (91.3% - 99.3%) | IgG:<br>25% (23.2% - 26.9%)<br><br>IgM:<br>0.2% (0.1% - 0.4%)<br><br>Combined IgG/IgM:<br>27.4% (25.6% - 29.3%) | (*19*) |
| Khartoum, Sudan | Survey of historic symptomatic infections conducted through social-media channels.<br><br>26/05/2020 - 03/06/2020 | (*25*) | Cross-sectional household study of 2375 people. 01/03/2021 - 10/04/2021 | STANDARD Q COVID-19 IgM/IgG Combo rapid diagnostic test (RDT) (SD–Biosensor).<br><br>ELISA (Anti–SARS-CoV-2 ELISA [IgG, S1 domain]; Euroimmun) conducted on subsample to estimate study-specific sensitivity and specificity using a meta-analysis of test characteristics with random effects and a Bayesian latent class model.<br><br>IgG: Sensitivity 61.9% (95% CI: 56.8–66.9)<br>IgG: Specificity of 98.9% (95% CI: 98.2–99.5). | Combined IgG/IgM (corrected for sensitivity/specificity): 54.6% (51.4% - 57.8%) | (*7*) |

**Table S2: p-values from Chi-squared test (df = 1) for difference between seroprevalence reportedby Abdella et al.** (*12*) **and estimated seroprevalence in Addis Ababa modelled under different mortality time series for different antibody types.** A p-value >0.05 indicates no statistically significant difference at the 0.05 level and is underlined below. Abdella et al. reported both unweighted and weighted estimates of seroprevalence from their study. The unweighted estimates are greater than the corresponding weighted estimates and in general showed a weaker agreement with modelled seroprevalence.

| | | **Excess mortality scenarios** | | | |
|---|---|---|---|---|---|
| | **COVID-19 deaths** | **2019 baseline** | **2019 baseline without 1st peak** | **2015 - 2019 baseline** | **2015 - 2019 baseline without 1st peak** |
| **Comparison to weighted seroprevalence reported by Abdella et al.** (*12*) | | | | | |
| **IgG** | 0.884 | <0.001 | 0.072 | 0.102 | 0.474 |
| **Combined IgG/IgM** | 0.334 | 0.002 | 0.347 | 0.765 | 0.074 |
| **Comparison to unweighted seroprevalence reported by Abdella et al.** (*12*) | | | | | |
| **IgG** | 0.112 | <0.001 | 0.487 | 0.771 | 0.007 |
| **Combined IgG/IgM** | 0.007 | 0.014 | 0.925 | 0.282 | <0.001 |

**Table S3: p-values from Chi-squared test (df = 1) for difference between seroprevalence observed by Bin-Ghouth et al. (*19*) and estimated seroprevalence in Aden under model fitted to satellite-derived excess mortality, modelled under different assumptions of the IFR and seroreversion half-life.** A p-value >0.05 indicates no statistically significant difference at the 0.05 level and is shown underlined.

| | | **IFR** | | | |
|---|---|---|---|---|---|
| | | **0.2** | **0.3** | **0.4** | **0.5** |
| **IgG** | | | | | |
| **IgG seroreversion half-life (days)** | **100** | <0.001 | <0.001 | <0.001 | <0.001 |
| | **120** | <0.001 | <0.001 | <0.001 | <0.001 |
| | **140** | <0.001 | <0.001 | <0.001 | <0.001 |
| | **160** | 0.155 | <0.001 | <0.001 | <0.001 |
| | **180** | 0.007 | 0.651 | 0.068 | <0.001 |
| | **200** | <0.001 | 0.001 | 0.289 | 0.089 |
| | **220** | <0.001 | <0.001 | 0.001 | 0.649 |
| | **240** | <0.001 | <0.001 | <0.001 | 0.005 |
| | **260** | <0.001 | <0.001 | <0.001 | <0.001 |
| | **280** | <0.001 | <0.001 | <0.001 | <0.001 |
| **Combined IgG/IgM** | | | | | |
| **IgG seroreversion half-life (days) with IgM half-life held constant at 50 days** | **100** | <0.001 | <0.001 | <0.001 | <0.001 |
| | **120** | <0.001 | <0.001 | <0.001 | <0.001 |
| | **140** | <0.001 | <0.001 | <0.001 | <0.001 |
| | **160** | 0.014 | <0.001 | <0.001 | <0.001 |
| | **180** | 0.089 | 0.563 | 0.003 | <0.001 |
| | **200** | <0.001 | 0.015 | 0.944 | 0.002 |
| | **220** | <0.001 | <0.001 | 0.031 | 0.385 |
| | **240** | <0.001 | <0.001 | <0.001 | 0.130 |
| | **260** | <0.001 | <0.001 | <0.001 | 0.005 |
| | **280** | <0.001 | <0.001 | <0.001 | <0.001 |
| **IgM** | | | | | |
| **IgM** | **30** | 0.012 | 0.019 | 0.025 | 0.026 |

| | | | | | |
|---|---|---|---|---|---|
| **seroreversion half-life (days)** | **35** | 0.016 | 0.031 | 0.045 | 0.045 |
| | **40** | 0.023 | 0.060 | 0.090 | 0.089 |
| | **45** | 0.042 | 0.131 | 0.191 | 0.183 |
| | **50** | 0.103 | 0.307 | 0.409 | 0.376 |
| | **55** | 0.249 | 0.567 | 0.683 | 0.623 |
| | **60** | 0.668 | 0.990 | 0.932 | 0.975 |
| | **65** | 0.799 | 0.662 | 0.634 | 0.734 |
| | **70** | 0.288 | 0.334 | 0.351 | 0.439 |

**Table S4: p-values from Chi-squared test (df = 1) Khartoum for reporting fractions explored in Figure 3.** A p-value >0.05 indicates no statistically significant difference at the 0.05 level and is shown underlined.

| **Reporting Fraction** | **p-values from Chi-squared test (df = 1)** |
|---|---|
| 2% | <0.001 |
| 3% | <0.001 |
| 4% | <0.001 |
| 4.5% | 0.492 |
| 5% | 0.118 |
| 6% | <0.001 |
| 7% | <0.001 |
| 8% | <0.001 |
| 9% | <0.001 |
| 10% | <0.001 |

**Table S5: Estimated reporting fractions of COVID-19 deaths as a percentage of total estimated excess deaths using alternative sources in each setting.** Uncertainty in total estimated excess deaths in Addis Ababa comes from consideration of different mortality baselines, with the 100% upper estimate.

| **Setting** | **Time period (inclusive)** | **Total reported COVID-19 deaths** | **Total estimated COVID-19 deaths (corresponding to reporting fraction range)** | **Estimating reporting fraction** |
|---|---|---|---|---|
| Addis Ababa, Ethiopia* | April 2020 - October 2020 | 1064 | 1064 - 1549 | 69% - 100% |
| Aden, Yemen † | March 2020 - September 2020 | 34 | 424 - 4137 | 0.8% - 8.0% |
| Khartoum, Sudan§ | March 2020 - April 2021 | 938 | 15630 - 31270 | 3.0% - 6.0% |

* 100% upper limit chosen to reflect both the near agreement between the cemetery inferred excess mortality and reported COVID-19 deaths, and the non-significant difference between the seroprevalence inferred based on model fits to reported COVID-19 deaths and the reported seroprevalence. 69% lower limit reflects the excess mortality estimated based on the 2015 - 2019 scenario with the first peak in May included.

† Reporting fraction range represents the uncertainty in the satellite-inferred excess mortality (95% CI: 424 - 4137).

§ Reporting fraction range represents the range of reporting fractions for which the corresponding modelled 95% confidence interval for the cumulative proportion of symptomatic infections overlapped with the reported lower or upper bounds of 8.3% and 13.7%.

**Table S6: Estimated reporting fractions of COVID-19 deaths as a percentage of total WHO-estimated excess deaths.** WHO excess mortality estimates calculated using the sum of months in which the WHO estimated positive excess mortality.

| **Setting** | **Time period (inclusive)** | **Total reported COVID-19 deaths** | **Total estimated positive excess deaths** | **Estimated reporting fraction** |
|---|---|---|---|---|
| Ethiopia | April 2020 - October 2020 | 1464 | 16656 (95% CI: 5608 - 34095) | 8.8% (95% CI: 4.3% - 26.1%) |
| Yemen | March 2020 - September 2020 | 588 | 10409 (95% CI: 6350 - 16748) | 5.6% (95% CI: 3.5% - 9.3%) |
| Sudan | March 2020 - April 2021 | 2080 | 20723 (95% CI: 12654 - 32395) | 10.0% (95% CI: 6.4% - 16.4%) |

#### Supplementary References


1. R. McCabe, O. J. Watson, *mrc-ide/covid-alternative-mortality: v0.1.0* (2022; https://zenodo.org/record/7185171).

2. P. G. T. Walker, C. Whittaker, O. J. Watson, M. Baguelin, P. Winskill, A. Hamlet, B. A. Djafaara, Z. Cucunubá, D. Olivera Mesa, W. Green, H. Thompson, S. Nayagam, K. E. C. Ainslie, S. Bhatia, S. Bhatt, A. Boonyasiri, O. Boyd, N. F. Brazeau, L. Cattarino, G. Cuomo-Dannenburg, A. Dighe, C. A. Donnelly, I. Dorigatti, S. L. van Elsland, R. FitzJohn, H. Fu, K. A. M. Gaythorpe, L. Geidelberg, N. Grassly, D. Haw, S. Hayes, W. Hinsley, N. Imai, D. Jorgensen, E. Knock, D. Laydon, S. Mishra, G. Nedjati-Gilani, L. C. Okell, H. J. Unwin, R. Verity, M. Vollmer, C. E. Walters, H. Wang, Y. Wang, X. Xi, D. G. Lalloo, N. M. Ferguson, A. C. Ghani, The impact of COVID-19 and strategies for mitigation and suppression in low- and middle-income countries. *Science*. **369**, 413–422 (2020).

3. O. J. Watson, M. Alhaffar, Z. Mehchy, C. Whittaker, Z. Akil, N. F. Brazeau, G. Cuomo-Dannenburg, A. Hamlet, H. A. Thompson, M. Baguelin, R. G. FitzJohn, E. Knock, J. A. Lees, L. K. Whittles, T. Mellan, P. Winskill, Imperial College COVID-19 Response Team, N. Howard, H. Clapham, F. Checchi, N. Ferguson, A. Ghani, E. Beals, P. Walker, Leveraging community mortality indicators to infer COVID-19 mortality and transmission dynamics in Damascus, Syria. *Nat. Commun.* **12**, 2394 (2021).

4. A. B. Hogan, P. Winskill, O. J. Watson, P. G. T. Walker, C. Whittaker, M. Baguelin, N. F. Brazeau, G. D. Charles, K. A. M. Gaythorpe, A. Hamlet, E. Knock, D. J. Laydon, J. A. Lees, A. Løchen, R. Verity, L. K. Whittles, F. Muhib, K. Hauck, N. M. Ferguson, A. C. Ghani, Within-country age-based prioritisation, global allocation, and public health impact of a vaccine against SARS-CoV-2: A mathematical modelling analysis. *Vaccine*. **39**, 2995–3006 (2021).

5. B. Borremans, A. Gamble, K. C. Prager, S. K. Helman, A. M. McClain, C. Cox, V. Savage, J. O. Lloyd-Smith, Quantifying antibody kinetics and RNA detection during early-phase SARS-CoV-2 infection by time since symptom onset. *Elife*. **9** (2020), doi:10.7554/eLife.60122.

6. N. F. Brazeau, R. Verity, S. Jenks, H. Fu, C. Whittaker, P. Winskill, I. Dorigatti, P. G. T. Walker, S. Riley, R. P. Schnekenberg, H. Hoeltgebaum, T. A. Mellan, S. Mishra, H. J. T. Unwin, O. J. Watson, Z. M. Cucunubá, M. Baguelin, L. Whittles, S. Bhatt, A. C. Ghani, N. M. Ferguson, L. C. Okell, Estimating the COVID-19 infection fatality ratio accounting for seroreversion using statistical modelling. *Commun. Med.* **2**, 54 (2022).

7. W. Moser, M. A. H. Fahal, E. Abualas, S. Bedri, M. T. Elsir, M. F. E. R. O. Mohamed, A. B. Mahmoud, A. I. I. Ahmad, M. A. Adam, S. Altalib, O. A. DafaAllah, S. A. Hmed, A. S. Azman, I. Ciglenecki, E. Gignoux, A. González, C. Mwongera, M. A. Miranda, SARS-CoV-2 Antibody Prevalence and Population-Based Death Rates, Greater Omdurman, Sudan. *Emerg. Infect. Dis.* **28**, 1026–1030 (2022).

8. EPHI Public Health Emergency Operations Center (PHEOC) COVID-19. *Ethiopian Public Health Institute*, (available at https://ephi.gov.et/download/ephi-pheoc-covid-19/).

9. Ethiopia: Country confirms first COVID-19-related death April 5 /update 7. *Crisis24* (2020), (available at https://crisis24.garda.com/alerts/2020/04/ethiopia-country-confirms-first-covid-19-related-death-april-5-update-7).

10. S. H. Watare, Z. A. Edea, H. T. Desta, F. Lombamo, N. Yohannes, M. Asefa, A. Mirkuzie, D. S. Gemechu, G. Teka, Y. D. Feyisa, K. Eshetu, M. Tesema, E. O. Musa, G. Tollera, A. Abayneh, E. Abate, The First Hundred Days of Novel Corona-virus Disease 2019 in Ethiopia, 13 March to 21 June 2020: Retrospective Analysis. *Research Square* (2020), doi:10.21203/rs.3.rs-74097/v1.

11. B. S. Endris, S. M. Saje, Z. T. Metaferia, B. G. Sisay, T. Afework, Y. G. Mengistu, E. H. Fenta, S. H. Gebreyesus, A. Petros, A. Worku, Y. Seman, D. H. Mariam, M. M. Sisay, A. Karim, Excess Mortality in the Face of COVID-19: Evidence from Addis Ababa Mortality Surveillance Program (2021), , doi:10.2139/ssrn.3787447.

12. S. Abdella, S. Riou, M. Tessema, A. Assefa, A. Seifu, A. Blachman, A. Abera, N. Moreno, F. Irarrazaval, G. Tollera, D. Browning, G. Tasew, Prevalence of SARS-CoV-2 in urban and rural Ethiopia: Randomized household serosurveys reveal level of spread during the first wave of the pandemic. *EClinicalMedicine*. **35**, 100880 (2021).

13. G. Timotewos, A. Solomon, A. Mathewos, A. Addissie, S. Bogale, T. Wondemagegnehu, A. Aynalem, B. Ayalnesh, H. Dagnechew, W. Bireda, E. S. Kroeber, R. Mikolajczyk, F. Bray, A. Jemal, E. J. Kantelhardt, First data from a population based cancer registry in Ethiopia. *Cancer Epidemiol.* **53**, 93–98 (2018).

14. United Nations Department of Economic and Social Affairs, World Urbanization Prospects: The 2018 Revision (2018), , doi:10.18356/b9e995fe-en.

15. A. D. Laytin, M. Sultan, F. Debebe, Y. Walelign, G. Fisseha, A. Gebreyesus, Critical care capacity in Addis Ababa, Ethiopia: A citywide survey of public hospitals. *J. Crit. Care*. **63**, 1–7 (2021).

16. S. Murthy, A. Leligdowicz, N. K. J. Adhikari, Intensive care unit capacity in low-income countries: a systematic review. *PLoS One*. **10**, e0116949 (2015).

17. Official account of Yemen Supreme National Emergency Committee for COVID-19. *Twitter*, (available at https://twitter.com/ysneccovid19).

18. E. S. Koum Besson, A. Norris, A. S. Bin Ghouth, T. Freemantle, M. Alhaffar, Y. Vazquez, C. Reeve, P. J. Curran, F. Checchi, Excess mortality during the COVID-19 pandemic: a geospatial and statistical analysis in Aden governorate, Yemen. *BMJ Glob Health*. **6** (2021), doi:10.1136/bmjgh-2020-004564.

19. A. S. Bin-Ghouth, S. Al-Shoteri, N. Mahmoud, A. Musani, N. M. Baoom, A. A. Al-Waleedi, E. Buliva, E. A. Aly, J. D. Naiene, R. Crestani, M. Senga, A. Barakat, L. Al-Ariqi, K. Z. Al-Sakkaf, A. Shaef, N. Thabit, A. Murshed, S. Omara, SARS-CoV-2 seroprevalence in Aden, Yemen: a population-based study. *Int. J. Infect. Dis.* **115**, 239–244 (2022).

20. A. A. Bawazir, Cancer incidence in Yemen from 1997 to 2011: a report from the Aden cancer registry. *BMC Cancer*. **18**, 540 (2018).

21. Department of Economic and Social Affairs, *World Population Prospects 2019 - Volume II: Demographic Profiles* (United Nations, 2020).

22. S. El Sirgany, S. Kiley, Coronavirus death rates in Yemen's Aden could exceed its wartime fatalities. *CNN* (2020), (available at https://edition.cnn.com/videos/tv/2020/06/10/ctw-yemen-covid-coronavirus.cnn).

23. حالة COVID-19 والتحديثات, (available at http://sho.gov.sd/corona/index.php).

24. تعميم إصابات و وفيات كوفيد-19 خلال... - وزارة الصحَّة الإتحاديَّة. *Federal Ministry of Health Sudan Facebook Page* (2020), (available at https://m.facebook.com/story.php?story_fbid=2764654530474174&id=182728912754405).

25. F. Mohamed, S. S. Satti, Maram, A. Ahmed, S. F. Babiker, Estimation of Coronavirus (COVID-19) Infections in Khartoum State (Sudan). *ResearchGate* (2020) (available at https://www.researchgate.net/publication/342122988_Estimation_of_Coronavirus_COVID-19_Infections_in_Khartoum_State_Sudan).

26. United Nations Office for the Coordination of Humanitarian Affairs, Sudan: Population Density and Potential COVID-19 Hotspots (2020), (available at https://reports.unocha.org/en/country/sudan/card/ZXsJqLs913/).

27. Sudan NextGen, "Statistics on Hospital Sector in Sudan" (2019), (available at https://sudannextgen.com/wp-content/uploads/2019/10/Statistics-on-Hospital-sector-in-Sudan-2.pdf).

28. H. Sulieman, W. El-Mahdi, M. Awadelkareem, L. Nazer, Characteristics of Critically-Ill Patients at Two Tertiary Care Hospitals in Sudan. *Sultan Qaboos Univ. Med. J.* **18**, e190–e195 (2018).

29. World Health Organization, "Methods for estimating the excess mortality associated with the COVID-19 pandemic" (2022), (available at https://www.who.int/publications/m/item/methods-for-estimating-the-excess-mortality-associatedwith-the-covid-19-pandemic).
